# Risk factors for severe respiratory syncytial virus infection during the first year of life: development and validation of a clinical prediction model

**DOI:** 10.1101/2023.02.23.23286237

**Authors:** Pekka Vartiainen, Sakari Jukarainen, Samuel Arthur Rhedin, Alexandra Prinz, Tuomo Hartonen, Andrius Vabalas, Essi Viippola, Rodosthenis S. Rodosthenous, Sara Kuitunen, Aoxing Liu, Cecilia Lundholm, Awad I. Smew, Emma Caffrey Osvald, Emmi Helle, Markus Perola, Catarina Almqvist, Santtu Heinonen, Andrea Ganna

**Affiliations:** Institute for Molecular Medicine Finland (FIMM), HiLIFE, University of Helsinki, Helsinki, Finland; Pediatric Research Center, Helsinki University Hospital and University of Helsinki, Helsinki, Finland; Department of Medical Epidemiology and Biostatistics, Karolinska Institutet, Stockholm, Sweden; Sachs’ Children and Youth Hospital, Stockholm, Sweden; Population Health Unit, Finnish Institute for Health and Welfare (THL), Helsinki, Finland; Broad Institute of MIT and Harvard, Cambridge, MA, USA; Pediatric Allergy and Pulmonology Unit at Astrid Lindgren Children’s Hospital, Karolinska University Hospital, Stockholm, Sweden; Stem Cells and Metabolism Research Program, Faculty of Medicine, University of Helsinki, Helsinki, Finland; Department of Paediatrics, Labatt Family Heart Centre, The Hospital for Sick Children, University of Toronto, Toronto, Ontario, Canada; Massachusetts General Hospital, Cambridge, MA, USA

## Abstract

**Background:** Novel immunisation methods against respiratory syncytial virus (RSV) are emerging, but knowledge of risk factors for severe RSV disease is insufficient for their optimal targeting. We aimed to identify predictors for RSV hospitalisation, and to develop and validate a clinical prediction model to guide RSV immunoprophylaxis for under 1-year-old infants.

**Methods:** In this retrospective cohort study using nationwide registries, we studied all infants born in 1997-2020 in Finland (n = 1 254 913) and in 2006-2020 in Sweden (n = 1 459 472), and their parents and siblings. We screened 1 510 candidate predictors and we created a logistic regression model with 16 predictors and compared its performance to a machine learning model (XGboost) using all 1 510 candidate predictors.

**Findings:** In addition to known predictors such as severe congenital heart defects (CHD, adjusted odds ratio (aOR) 2·89, 95% confidence interval 2·28-3·65), we identified novel predictors for RSVH, most notably esophageal malformations (aOR 3·11, 1·86-5·19) and lower complexity CHDs (aOR 1·43, 1·25-1·63).

In validation data from 2018-2020, the C-statistic was 0·766 (0·742-0·789) in Finland and 0·737 (0·710-0·762) in Sweden. The clinical prediction model’s performance was similar to the machine learning model (C-statistic in Finland 0·771, 0·754-0·788). Calibration varied according to epidemic intensity. Model performance was similar across different strata of parental income.

The infants in the 90th percentile of predicted RSVH probability hospitalisation had 3·3 times higher observed risk than the population’s average. Assuming 60% effectiveness, immunisation in this top 10% of infants at highest risk would have a number needed to treat of 23 in Finland and 40 in Sweden in preventing hospitalisations.

**Interpretation:** The identified predictors and the prediction model can be used in guiding RSV immunoprophylaxis in infants.

**Funding:** Sigrid Jusélius Foundation, European Research Council, Pediatric Research Foundation (for complete list of funding sources, see Acknowledgements).

## Introduction

Respiratory syncytial virus (RSV) causes respiratory infections across all ages, but the majority of disease burden affects young children.^1^ In infants less than 12 months old, severe lower respiratory tract infections due to RSV leading to hospitalisation (RSVH) have an estimated incidence from 15 to 35 per 1000 children.^2^,^3^ Over 100 000 deaths in under 5-year-old children can be attributed to RSV each year globally.1 RSV infections occur in annual epidemics during cold months in temperate climates, some countries having a biennial variation in this pattern,^4^–^6^ whereas in the tropics the epidemics are more variable.^7^ The RSV epidemic patterns were disturbed globally by the COVID-19 pandemic and infection control restrictions, with particularly strong and early epidemics occurring throughout the world during 2021-2022 after a hiatus.^8^,^9^

Use of the only immunoprophylaxis against RSV available, monoclonal antibody palivizumab, is restricted to a limited number of high-risk children due to high pricing and cumbersome dosing. Major advances have recently been made in developing novel immunisation methods against RSV.^10^ Nirsevimab,^11^ a long-acting monoclonal antibody that can be administered as a single injection inducing protection for a whole RSV epidemic, was recently approved by the European Medicines Agency. Also maternal vaccines against RSV acting through placental antibody transfer ^12^ have shown efficacy in clinical trials.

These novel immunoprophylaxis methods will likely be available to a larger population than palivizumab, but balancing costs and population health impact is crucial,^13,14^ making the identification of high-risk infants a key problem. Age is the most important risk factor for RSVH, as most hospitalisations occur in infants aged 6 months or less.^2,9^ Other well characterised predisposing medical conditions include preterm birth, bronchopulmonary dysplasia (BPD),^15^ and severe congenital heart defects (CHD),^16^ but most hospitalised children do not have any of these conditions. Several environmental and host-related factors associated with RSVH have been identified, including the number of young siblings, neonatal respiratory problems and tobacco smoke exposure.^17–19^ Still, the combined effect of different predictors of RSVH is poorly understood, and studies assessing and comparing a broad range of predictors are lacking. All existing prediction models of severe RSV disease have major limitations (see **supplementary material** for details), and current immunoprophylaxis guidelines are not based on systematic risk stratification and vary across countries.^20,21^

## Aim

In this study, we aimed to identify predictors for RSV bronchiolitis hospitalisation during the first year of life by analysing nationwide registry data and to develop a clinical prediction model to guide the passive RSV immunisation for under 1-year-old infants.

## Methods

### Study population and setting

See **Figure 1** for a graphical overview of the study setting and data. We examined all children born in Finland between June 1, 1997 and May 31, 2020, and their parents and siblings, using FinRegistry data (www.finregistry.fi). From Sweden, we examined all children born in Sweden between June 1, 2006 and May 31, 2020.

**Figure 1.**
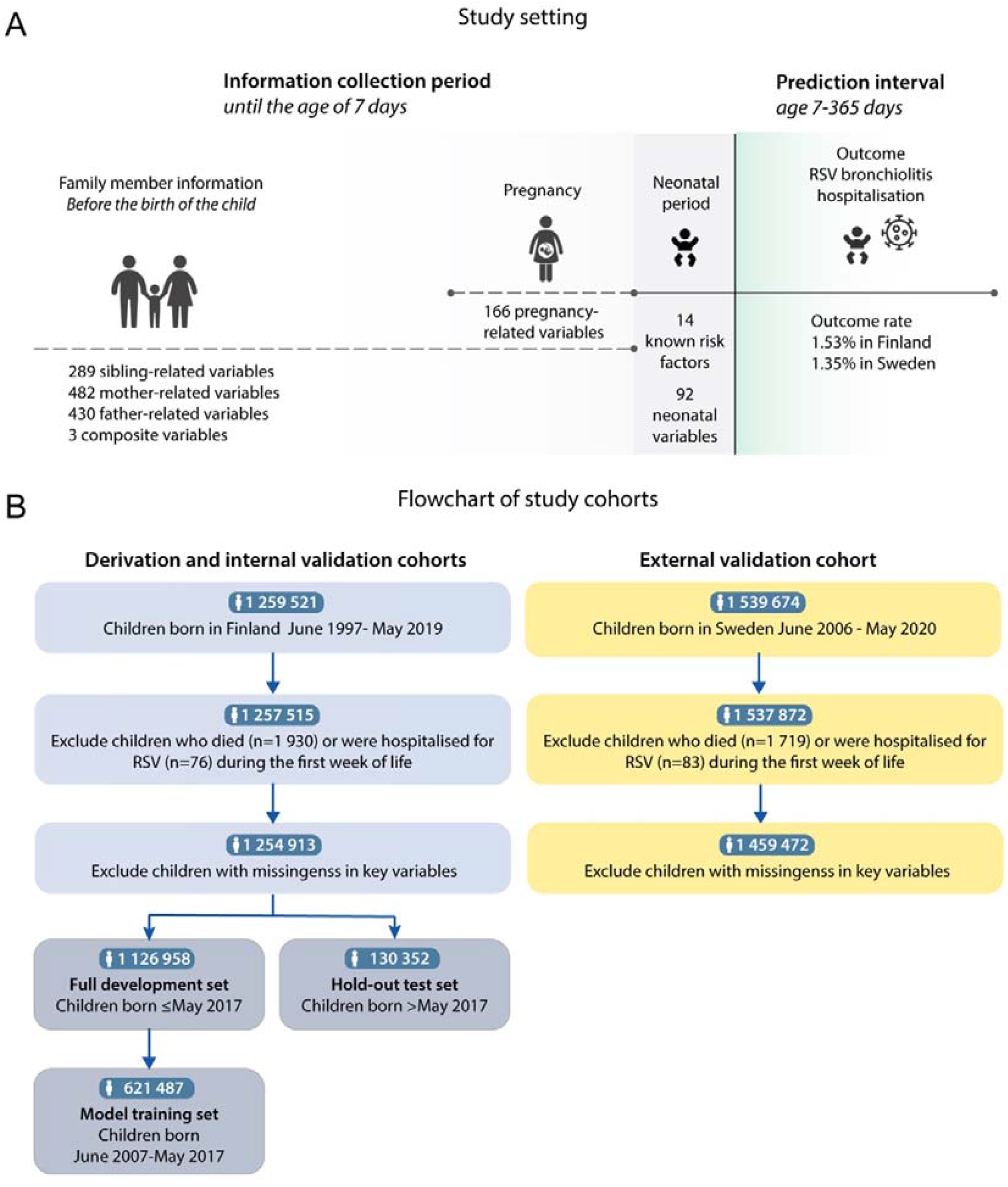
The study setting and study population summarised. Panel A shows the study setting as a timeline, illustrating the different sources of information used to define the predictors and the outcome. Panel B shows the study population definition as a flowchart.

The exclusion criteria were death or RSVH during 0-7 days of age. As missingness was rare, we performed the analyses using complete cases. In Finland, 2 602 (0·21%) with missingness were excluded from model development and validation. In Sweden, 78 400 (5·1%) were excluded from external validation because of missingness. The missing data are further detailed in **the supplementary material**.

The predicted outcome was hospitalisation with a diagnosis of RSV bronchiolitis (RSVH), defined as having an inpatient hospital episode with an ICD-10 diagnosis code of RSV bronchiolitis (J21.0) within 7-365 days of age (the prediction interval). In the Finnish cohort we also compared laboratory-confirmed RSV detections to RSVH cases (hospitalisation with ICD-10 code J21.0) as a sensitivity analysis of outcome validity. The children were followed until 1 year of age.

Because RSV epidemics often occur close to the change of the calendar year, we defined the RSV epidemic year as starting on June 1st and ending on May 31st.

### Predictors of RSV hospitalisation

#### Predefined predictors

First, we defined a set of 14 predefined predictors for RSVH based on the literature ^17,18,22–26^. Subsequent screening of other candidate predictors was always performed controlling for the effect of these 14 predefined predictors. The predefined predictors were 1) gestational age at birth (in days) 2) Months from birth to the next estimated epidemic peak, 3) mother’s age at birth (in years), 4) father’s age at birth (in years), 5) twinhood; 6) older siblings aged <4 years; 7) older siblings aged 4-7 years; 8) older siblings over 7 years; 9) birth weight in SD units, 10) Down syndrome; 11) BPD; and 12-14) severe CHD requiring operation during the first month, the first year of life or later in life. See **supplementary material** for their definitions. The continuous variables were modelled with restricted cubic splines with 4-6 knots.

#### Candidate predictors

We considered altogether 1 510 candidate predictors: 14 predefined, 487 mother-related, 434 father-related, 314 sibling-related, 166 pregnancy-specific and 92 infant-related predictors, and 3 composite variables (e.g. asthma in any first-degree relative). For the infant, we considered information on neonatal period, congenital conditions and birth-related variables from the Medical Birth Register, the Congenital Malformations Register, and the hospital registries prior to the age of 7 days. For mothers we defined pregnancy-specific variables from diagnoses, drug purchases and pregnancy-related registry variables. Family members’ predictors included medical events (using pre-existing composite disease definitions from the FinnGen study), and drug purchases until the birth of the child. See supplementary material for details of the variable definitions.

#### Predictor discovery and model development

We followed the TRIPOD recommendations for clinical prediction model development and reporting.^27^ The Finnish study population was divided into a development dataset (birth between June 1997 and May 2017, n = 1 126 952) and an temporal hold-out validation dataset (birth between June 2017 and May 2020, n = 130 352). The full development dataset was used for predictor screening, but the last 10 years were used to estimate the final model coefficients (birth between June 2006 and May 2017, n = 621 487) to maximise generalisation to future years. External validation was performed in the Swedish cohort (n = 1 459 472).

For detailed description of the methods, please see supplementary material (subheading “Predictor selection”). First, we screened for preliminary associations between the candidate predictors and RSVH with multiple logistic regression models adjusted for the 14 predefined predictors. After this screening, we combined the 14 predefined predictors (known risk factors) and the most important candidate predictors to a single multivariable logistic model, and considered results from backwards stepwise elimination using the Akaike information criterion (AIC) and L1-regularised (Lasso) logistic regression.

Finally, we removed, modified and combined predictors based on expert judgement, making trade-offs between predictive performance, simplicity, ease of application in the clinical context and expected generalisability. This rationale is also described in more detail in the supplementary material.

After selecting the predictors, the final prediction model was trained with logistic regression. To describe the relative importance of predictors in population level, we compared their population attributable fractions.^28^

#### Model validation

We assessed the model discrimination (how well the predictions differentiate infants with an outcome from those without an outcome) with the C-statistic. Calibration (the agreement between predicted and observed risk) was assessed graphically in deciles divided by the predicted probability, and by calculating the calibration slope and the intercept.

Discrimination and calibration were estimated separately for each epidemic year of the validation data and pooled estimates were obtained using random effects meta-analysis.

In parallel with the logistic regression model using 16 predictors, we developed a complex machine learning model with eXtreme Gradient Boosting (XGBoost, ^29^) using all defined 1 510 candidate predictors. This model was only tested in the Finnish validation data. SHapley Additive exPlanation (SHAP) values were used to assess variable importance in the XGBoost model To assess the potential clinical utility of the model, we first performed a decision curve analysis.^30^ Second, we estimated a hypothetical number needed to treat (NNT) value to prevent one RSVH if an immunpoprophylaxis were targeted using the percentile cutoffs of the predicted probability from the model. We assumed conservative 60% efficacy of the immunoprophylaxis in preventing hospitalisations.^11,31^ To study the model’s fairness, we analysed the C-statistic of the model’s predictions across parental income quintiles. See supplementary material for more details of the model validation methods.

#### Ethical considerations

FinRegistry is a joint project of the Finnish Institute for Health and Welfare (THL) and the Data Science Genetic Epidemiology research group at the Institute for Molecular Medicine Finland (FIMM), University of Helsinki. The FinRegistry project has received the following approvals for data access from the National Institute of Health and Welfare (THL/1776/6.02.00/2019 and subsequent amendments), Digital and population data services agency DVV (VRK/5722/2019-2), Finnish Center for Pension (ETK/SUTI 22003) and Statistics Finland (TK-53-1451-19). The FinRegistry project has received IRB approval from the National Institute of Health and Welfare (Kokous 7/2019).The study was also approved by the Swedish Ethical Review Authority (Dnr 2018/1697-31/1 and Dnr 2022-05010-02). Prior to starting the analyses, we archived a study protocol.^32^

## Results

The characteristics of the 2·72 million included children are presented in **table 1**.

**Table 1.**
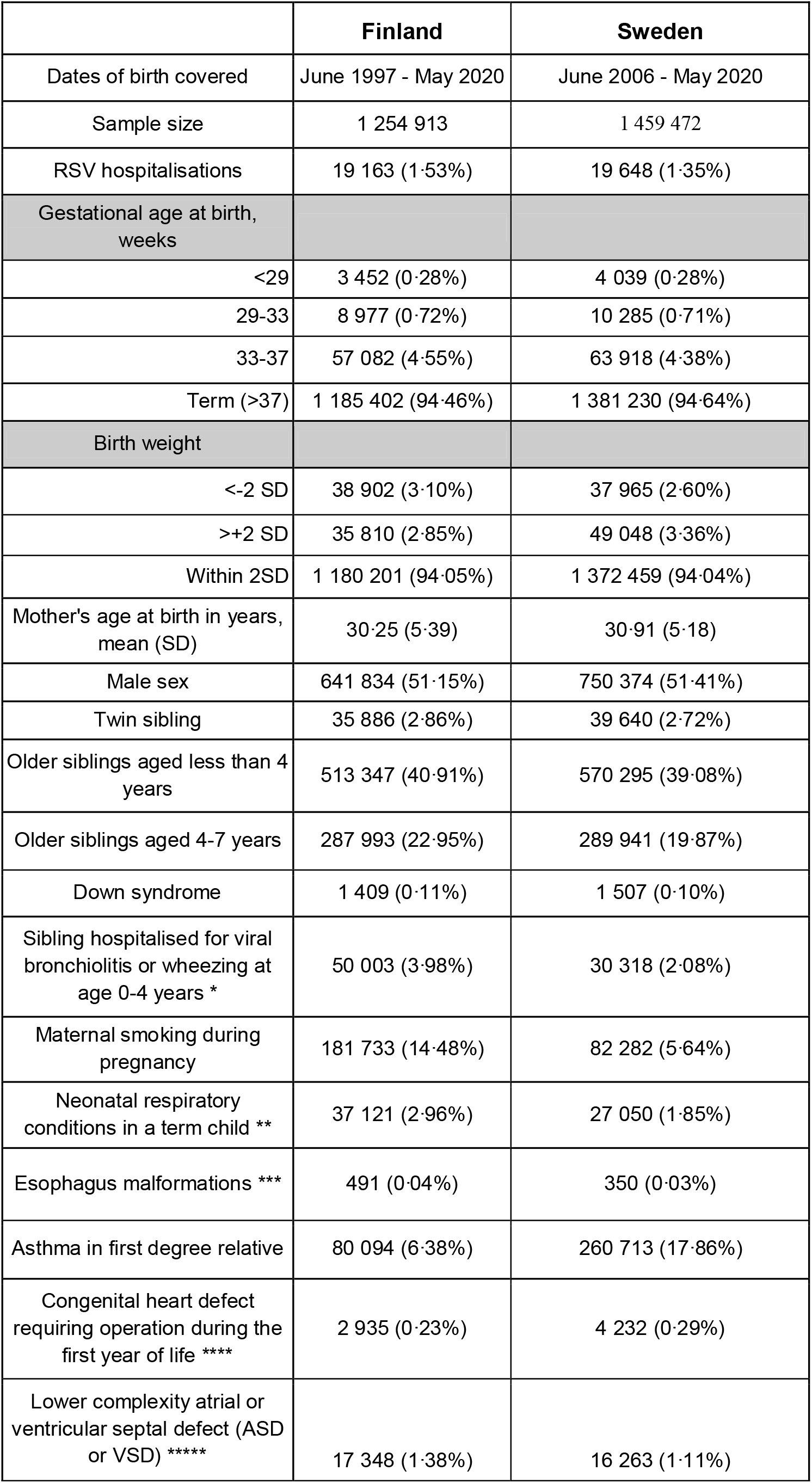
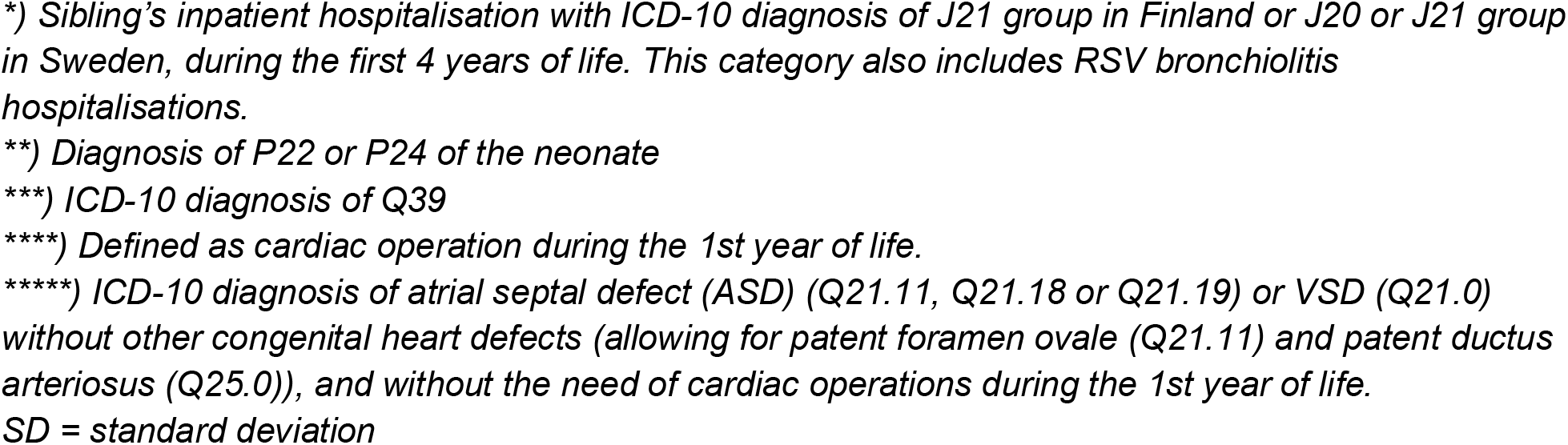
The characteristics of the children included in the study, shown for the variables of the final prediction model. *) Sibling’s inpatient hospitalisation with ICD-10 diagnosis of J21 group in Finland or J20 or J21 group in Sweden, during the first 4 years of life. This category also includes RSV bronchiolitis hospitalisations. **) Diagnosis of P22 or P24 of the neonate ***) ICD-10 diagnosis of Q39 ****) Defined as cardiac operation during the 1st year of life. *****) ICD-10 diagnosis of atrial septal defect (ASD) (Q21.11, Q21.18 or Q21.19) or VSD (Q21.0) without other congenital heart defects (allowing for patent foramen ovale (Q21.11) and patent ductus arteriosus (Q25.0)), and without the need of cardiac operations during the 1st year of life. SD = standard deviation

|  | Finland | Sweden |
| --- | --- | --- |
| Dates of birth covered | June 1997 - May 2020 | June 2006 - May 2020 |
| Sample size | 1 254 913 | 1 459 472 |
| RSV hospitalisations | 19 163 (1.53%) | 19 648 (1.35%) |
| Gestational age at birth, weeks |  |  |
| <29 | 3 452 (0.28%) | 4 039 (0.28%) |
| 29-33 | 8 977 (0.72%) | 10 285 (0.71%) |
| 33-37 | 57 082 (4.55%) | 63 918 (4.38%) |
| Term (>37) | 1 185 402 (94.46%) | 1 381 230 (94.64%) |
| Birth weight |  |  |
| <-2 SD | 38 902 (3.10%) | 37 965 (2.60%) |
| >+2 SD | 35 810 (2.85%) | 49 048 (3.36%) |
| Within 2SD | 1 180 201 (94.05%) | 1 372 459 (94.04%) |
| Mother's age at birth in years, mean (SD) | 30.25 (5.39) | 30.91 (5.18) |
| Male sex | 641 834 (51.15%) | 750 374 (51.41%) |
| Twin sibling | 35 886 (2.86%) | 39 640 (2.72%) |
| Older siblings aged less than 4 years | 513 347 (40.91%) | 570 295 (39.08%) |
| Older siblings aged 4-7 years | 287 993 (22.95%) | 289 941 (19.87%) |
| Down syndrome | 1 409 (0.11%) | 1 507 (0.10%) |
| Sibling hospitalised for viral bronchiolitis or wheezing at age 0-4 years * | 50 003 (3.98%) | 30 318 (2.08%) |
| Maternal smoking during pregnancy | 181 733 (14.48%) | 82 282 (5.64%) |
| Neonatal respiratory conditions in a term child ** | 37 121 (2.96%) | 27 050 (1.85%) |
| Esophagus malformations *** | 491 (0.04%) | 350 (0.03%) |
| Asthma in first degree relative | 80 094 (6.38%) | 260 713 (17.86%) |
| Congenital heart defect requiring operation during the first year of life **** | 2 935 (0.23%) | 4 232 (0.29%) |
| Lower complexity atrial or ventricular septal defect (ASD or VSD) ***** | 17 348 (1.38%) | 16 263 (1.11%) |

\*\*) Diagnosis of P22 or P24 of the neonate
\*\*\*) ICD-10 diagnosis of Q39
SD = standard deviation

### Incidence of RSV hospitalisations

During the first year of life 19 195 children (1·53 %) in the Finnish and 19 648 (1·35%) in the Swedish cohort had RSVH, most often during the first months of life (**supplementary figure 2**). In the Finnish data, 83·6% of children with RSVH had a recorded laboratory confirmed RSV infection within 7 days of hospitalisation in 2007-2020, with a higher rate in more recent years (93·1% in 2018-2020), and the seasonality of RSVH and positive RSVH tests in Finnish National Infectious Diseases Register was similar (**supplementary figures 2** and **3)**.

### Predictor discovery

Most of the 14 predefined predictors based on the literature were significantly associated with RSVH (**supplementary table 1**). BPD did not have an independent effect (aOR 0·74, 95% CI 0·59-0·93).

We systematically screened for new predictors by testing the association between 1496 candidate predictors from parents, siblings and the infant. **Figure 2** illustrates the odds ratios adjusted for predefined predictors (aOR) between the candidate predictors and RSVH. In **supplementary results**, more associations are detailed. Complete list of estimates by predictor category are presented in **supplementary tables 2-7**.

**Figure 2.**
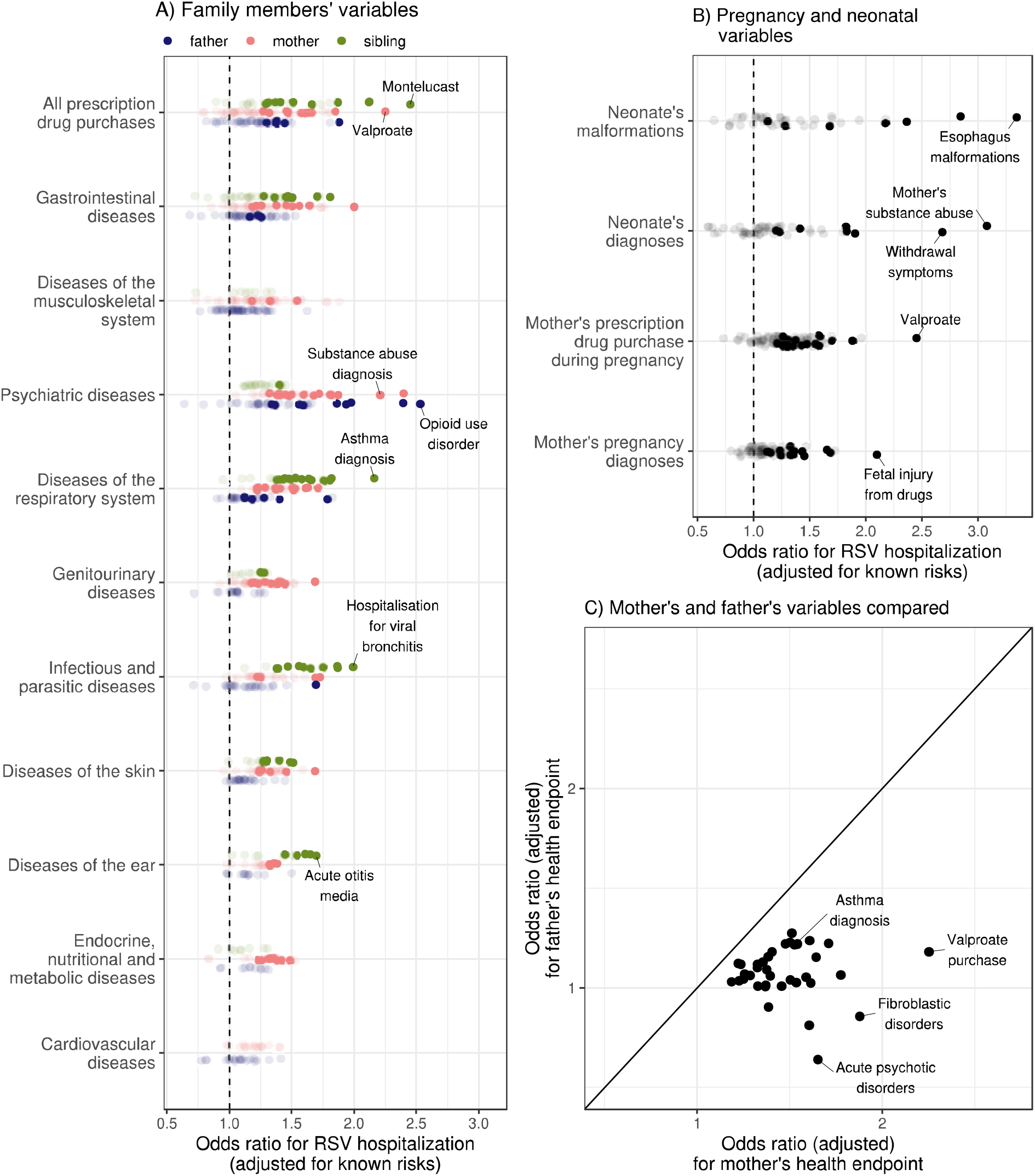
The odds ratios adjusted for the 14 predefined predictors (aOR) obtained from predictor screening of diagnoses and drug purchases of the infant and family members. **Panel A** shows the comparison of the aORs of parents’ and siblings’ health events. Colours indicate different family members’ variables. **Panel B** shows the aORs for pregnancy and neonatal variables. Solid points indicate statistically significant aORs after correction for multiple testing. For family members’ health events, the threshold for the multiple testing-corrected p value is p<6e-5 but for neonatal conditions a lower threshold of p<1e-3 was used because of the lower number of conditions included in these categories. **Panel C** shows the comparison of mothers’ and fathers’ aORs. Only disease conditions which have indication of statistically significant difference between mothers and fathers in their regression coefficients (p<0·01 in Z-test, not corrected for multiple testing), are compared. The points below the diagonal reference line indicate that aORs were higher in mothers.

**Figure 3.**
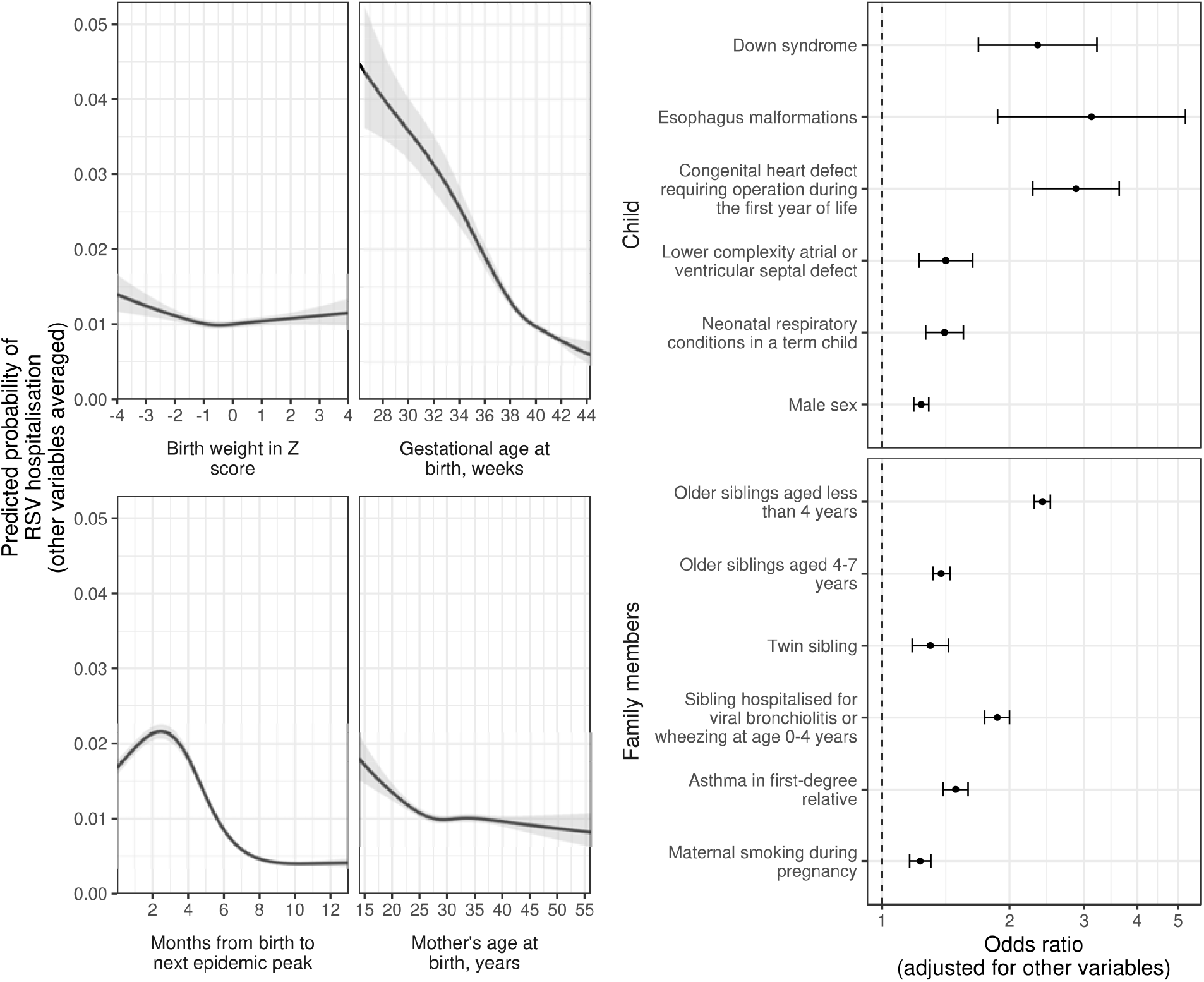
The final prediction model variables and their individual association with the probability of RSV hospitalisation. For continuous variables, the predicted probability and its 95% confidence interval is shown for the range of the variable, when the other variables in the model are averaged. For binary variables (right), odds ratios adjusted for other variables in the model are shown.

Among infants’ diagnoses, esophagus malformation (aOR 3·34, 95% CI 2·13-5·25) had the highest effect size. Lower complexity CHDs increased the risk of RSVH. Uncomplicated atrial septal defect (ASD), i.e. diagnosis of ASD without heart operations or other CHD diagnoses (aOR 1·68, 95% CI 1·42-1·98) and uncomplicated ventricular septal defect (VSD, aOR 1·28, 95% CI 1·11-1·48) were both associated with increased risk. For the final prediction model we combined them into a composite variable (having lower complexity ASD or VSD, aOR 1·41, 95% CI 1·25-1·60).

Among the parents’ prior diagnoses, psychiatric diseases and substance use disorder diagnoses, both in mothers and fathers, were clearly associated with increased RSVH risk (**figure 1a**). Especially substance abuse diagnoses, both from time before pregnancy and during pregnancy, were strongly associated (**Figure 1, supplementary results**). Overall, the associations between the parents’ health events and RSVH in the infant were stronger in mothers than fathers (**figure 1c**).

### Clinical prediction model

Of the 1 510 variables screened, we selected 16 predictors that are easily ascertained in a clinical context and plausibly generalise to other countries. The variables and their independent association with RSVH are shown in **figure 2**. The prediction model is presented as an online calculator, available at https://rsv-risk-1.netlify.app/. The variable definitions, as well as their regression coefficients are presented in supplementary table 8. The distribution of the model’s predicted probabilities was similar across different years (**supplementary figure 4**). **Supplementary figure 5** shows the population attributable fractions of the predictors.

### Model discrimination and calibration

The meta-analysed C-statistic from each individual epidemic year of the Finnish training data (epidemic years 2007-2017) was 0·744 (95% CI 0·734-0·754). **Supplementary figures 6 and 7** show the discrimination and calibration measures in the training data, compared to those from external validation data from Sweden. In the complete pooled Swedish validation data, the C-statistic was 0·729 (95% CI 0·725-0·732).

**Figure 4**. shows the comparison of discrimination and calibration measures in the 3 most recent epidemics (2018-2020) of the validation data. The C-statistics in Sweden were 0·737 (95% CI 0·710 - 0·762) using meta-analysis, and 0·741 (95% CI 0·733-0·749) using pooled data. The respective C-statistics in the Finland were (0·766, 95% CI 0·742-0·789) using meta-analysis and 0·767 (95% CI 0·759-0·775) using pooled data.

**Figure 4.**
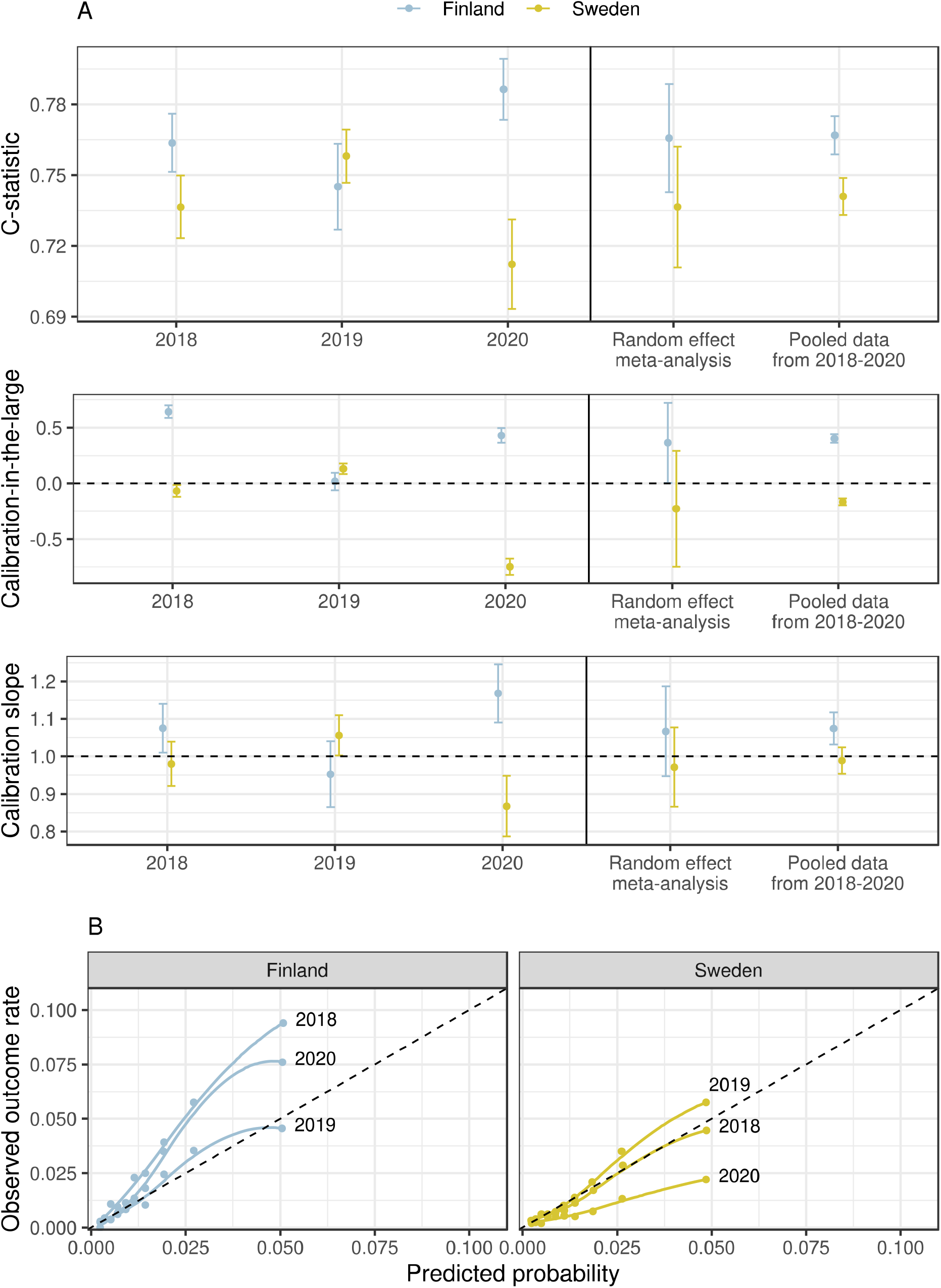
The discrimination and calibration of the prediction model in the hold-out validation datasets from Finland and Sweden. Panel A shows the model performance metrics individually for each RSV epidemic year in 2018-2020, the random effects meta-analysis from those years, and finally pooling the data from those three epidemic years. Panel B shows the calibration plots, i.e. the predicted vs the observed probabilities in equally sized deciles divided according to the predicted probability,separately for the three epidemic years of this comparison. The dashed reference lines indicate perfect calibration. There was notable variation in the calibration during individual epidemic years: too low predicted probabilities occurred in strong epidemics and too high in mild epidemics.

We observed variation in model calibration corresponding to variation in RSV epidemic intensity. Too low predicted probabilities occurred in strong epidemics and too high in mild epidemics (**figure 4, supplementary figures 6, 7** and **8**), nonetheless the average calibration was good in internal and external validation. We also observed some variation in model discrimination, strong epidemics having slightly better C-statistic than milder ones (**figure 4** and **supplementary figure 7**).

### Secondary analyses of model performance

The C-statistic of the XGboost model containing all 1 510 predictors was 0·771 (95% CI 0·754 - 0·788) in the Finnish hold-out validation data, representing a very limited increase in discrimination compared to the main model (C-statistic of 0·766, 95% CI 0·742 - 0·789).

Calibration showed a similar trend of higher predicted probabilities in the two strong epidemics included to the Finnish hold-out validation data. The full results of this comparison are shown in **supplementary figure 9. Supplementary figure 10** shows the most important variables in the XGboost model.

In the decision curve analysis, the clinical prediction model showed higher net benefit than treat all, treat none, and the AAP criteria strategies for a wide range of threshold probabilities of up to approximately 0·1 in Finland and up to 0·075 in Sweden (**figure 5a**).

**Figure 5.**
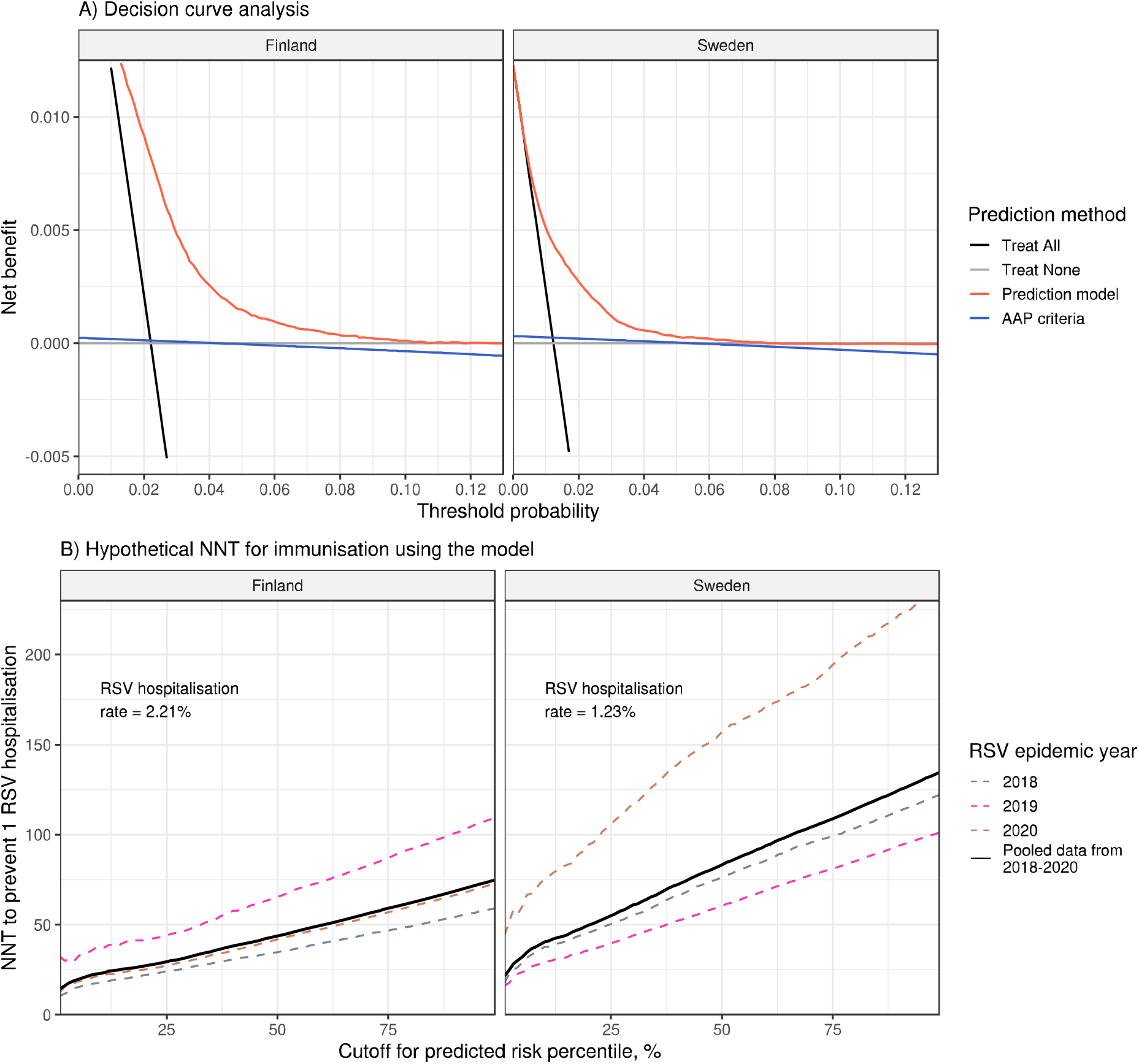
Model utility analyses in the hold-out validation data of children born between 6/2017-5/2020. **Panel A** shows the decision curve analysis using Finnish and Swedish hold-out validation data. The y-axis shows the net benefit, i.e. the trade-off between true positives and false positives weighed for the cutoff probability for the range of threshold probability 0 - 0·15. Four intervention strategies are compared: treat all, treat none, treat according to the American Academy of Pediatrics (AAP) palivizumab prophylaxis guidelines ^20^ and treat according to the presented prediction model. Based on this analysis, using the prediction model to target immunoprophylaxis would be beneficial over the other strategies if the benefits of immunoprophylaxis outweigh the harms for infants with probability of RSV hospitalisation of 0·1 or less (0·075 or less in Sweden). The AAP criteria were inferior for all analysed thresholds. **Panel B** shows the hypothetical number-needed to treat (NNT) estimates for targeting immunoprophylaxis according to the percentiles of the prediction model’s predicted risk. Y-axis shows the NNT for immunoprophylaxis, assuming 60% efficacy in preventing RSV hospitalisations. X-axis shows the cutoff percentile for immunisation. For example, in the Finnish validation data, the top 10% infants with highest predicted risk of RSVH, the observed RSVH risk was 3·3 times higher (7·3%, vs 2·2%) than in all infants during the 2018-2020 epidemics, and the RSVHs in this top 10% accounted for 33% of all RSVHs. The NNT in this top 10% highest risk group of infants would be 23 (ranging from 18 in 2018 to 36 in 2019), and the prophylaxis would have prevented 20% of all RSVHs (ranging from 18% in 2019 to 21% in 2018). Similarly in Sweden the top 10% highest predicted risk infants had 3·3 times higher observed risk of RSVH than all infants (4·2% vs 1·3%). The RSVHs in these top 10% accounted for 32% of all RSVHs. The NNT would have been 40 (ranging from 29 in 2019 to 77 in 2020), and the prophylaxis would have prevented 19% of all RSVHs (ranging from 18% in 2020 to 20% in 2019).

To further evaluate the potential clinical utility of the model we estimated the NNT to prevent one RSVH for an immunoprophylaxis targeted according to the prediction model risk percentiles in the validation data (**Figure 5b**). Despite varying NNT estimates according to the outcome prevalence in different epidemics, NNT estimates showed a clear increasing trend corresponding to increasing cutoff percentile. See **supplementary results** for further examples of the NNT calculations.

We tested the model’s fairness by estimating the C-statistic stratified by parental income quintiles in the Finnish hold-out data (**supplementary figure 11**). The C-statistic in the lowest quintile was 0·777 (95% CI 0·760-0·794), compared to 0·753 (0·731-0·775) in the highest quintile.

## Discussion

The recent surge in RSV infections following the COVID-19 pandemic and the upcoming new immunoprophylaxis methods highlight the need to identify infants at increased risk for severe RSV lower respiratory tract infection. In this study, we screened 1 510 candidate predictors for hospitalisation with RSV bronchiolitis during the first year of life (RSVH), covering the infant’s diagnoses, pregnancy and birth information and extensive disease and medication information of parents and siblings. We present a simple clinical prediction model for RSVH risk during the first year of life with satisfactory predictive performance in validation across different RSV epidemics in two countries. The model showed potential clinical utility in decision curve analysis and NNT simulations, and its performance was fair across different strata of parental income.

### Predictors for RSV hospitalisation

While we confirmed associations of several previously identified risk factors, we also identified new predictors for the risk of RSVH.

Among known risk factors, months from birth to the next estimated epidemic peak, gestational age at birth and having young siblings were the most important predictors of RSVH at the population level. For example, being 2 months old at the epidemic peak was associated with 5·3-fold risk of RSVH compared to age of 11 months, and being born before 29 weeks of gestation was associated with 4·2-fold risk compared to term children, other variables held equal. Having young siblings (aOR 2·42) also markedly increased RSVH risk, likely mediated through increased likelihood of RSV transmission to the infant.^24^

Hemodynamically significant CHDs are consistently recognised as a major risk factor for RSVH,^16^ but in this study, we identified lower complexity CHDs (uncomplicated ASD and VSD) as predictors for RSVH, of which there is only preliminary evidence.^33^ Lower complexity CHDs and similar, more common but less pronounced risk factors may contribute significantly to the population burden of RSVH, and are likely important to be considered in RSV immunisation targeting if it will be available for more broad populations.

Bronchopulmonary dysplasia, despite being perhaps the most established risk factor for RSVH, did not have an independent effect in the present models. This is probably a result of including gestational age in the model, which is strongly associated with BPD risk, and due to use of palivizumab prophylaxis in infants with BPD.

Among less established predictors, esophageal malformations increase the risk of hospitalisation due to respiratory infections and pneumonia during the first year of life,^34^ and a significant fraction of these cases have been attributed to RSV.^35^ Also Down syndrome has been shown to be a strong predisposing condition for RSVH.^23^ Neither Down syndrome nor esophageal malformations are currently considered as an indication for palivizumab prophylaxis in the current guidelines,^20,21^ but our results suggest that they are among the most significant underlying conditions associated with RSVH.

Family members’ mental health-related conditions and family income were consistently associated with RSVH, similarly than in a Canadian population-based study showing associations between adverse socioeconomic factors and RSVH.^36^ Because of the potential challenges related to generalisability and consistent ascertainment of predictors in clinical context, we decided not to include these predictors in the final prediction model. Moreover, parent’s mental health-related conditions might be associated with higher RSVH risk through mechanisms other than actual increased risk of severe RSV in the infant.

Family members’ asthma-related variables were consistently associated with RSVH. We could also quantify the differences in association between family members’ asthma-related variables and infant’s RSVH, with older sibling’s montelucast medication and previous hospitalisation with wheezing or bronchiolitis having the strongest associations. History of asthma in the mother was more strongly associated with RSVH risk than in the father, similarly to results of an interview-based case-control study.^37^ Also more generally, the same diagnoses in mothers were more predictive for RSVH than in fathers. In addition to possible biological mechanisms (such as breastfeeding), this might reflect differences in societal norms concerning childcare, with mothers usually being more responsible for the care of newborn children.

### Clinical prediction model for the risk of RSVH

Currently no widely used tool for predicting the risk of a severe RSV disease in the general infant population exists. We present a model using 16 easily ascertainable predictors, and having similar predictive performance to a more complex machine learning model (XGBoost) utilising 1 510 predictors. The model showed satisfactory performance in validation data from Finland and Sweden, containing epidemics with considerably varying outcome rates.

Moreover, the model appeared not to harmfully discriminate against children from lower income families. The timing of the annual RSV epidemics is an important determinant for the risk of RSVH in infancy. The present model requires estimating the timing of the next epidemic peak. We used a method of averaging peak months two, four and six years prior to the infant’s birth because of the biennial pattern observed in the Nordics,^5,6^ but any estimate based on best available knowledge can be used. The proposed model allows immunisation planning before the start of the RSV season is known, which is essential to achieve timely RSV immunisation for broader populations, and all validation analyses were done restricting to epidemiologic data available immediately after the birth of the infant.

The varying incidence of RSV infections across years and regions poses challenges to model calibration, especially after the Covid-19 pandemic. Although, on average, the model was similarly well calibrated in both Finland and Sweden, the observed risk during individual strong epidemics was close to 2-fold compared to the risk predicted by the model. While the prediction model can be recalibrated to other countries based on historical information on incidence of RSV infections, there always remains considerable uncertainty in the strength of future RSV epidemics and consequently well-calibrated predictions are difficult to achieve.

Instead, we have shown that epidemic intensity does not have a considerable impact on model discrimination (the ability to correctly rank children relative to each other according to their risk), and that this satisfactory discrimination translated into potential clinical utility in two countries, in varying intensities of epidemics and in various cutoff thresholds.

### Strengths, limitations and future research

Perhaps the most notable strength of this study lies in the wide variety of nationwide data available from two countries across multiple years. This allowed us, for example, to screen all family member’s diagnoses and medication purchases for potential predictors, to evaluate the joint effect of a wide range of predictors, and to create a complex machine learning model as a comparison for the logistic regression model. We have made extensive efforts considering multiple statistical approaches and clinical rationale to extract easily definable predictors to the clinical prediction model.

In our study we could not control for the effect of existing palivizumab prophylaxis. However, the use of palivizumab in Finland and Sweden is very conservative compared to e.g. AAP guidelines ^20^. Based on the palivizumab consumption statistics obtained from the Finnish Medical Agency we estimate that annually 100-200 Finnish children have received palivizumab prophylaxis. This is likely not affecting the results outside infants with BPD. Some rare conditions, or diseases that are diagnosed later during infancy, could not be captured by the present approach, but these conditions likely have extremely small impact to population-level prediction. Also some potentially relevant variables, such as breastfeeding, could not be included. Finally, conclusions about causality of the associated predictors and RSVH cannot be drawn based on this study.

It remains to be assessed how well the model generalises to countries with different RSV epidemics and health care systems, and the performance and fairness of the model should be studied in ethnically diverse populations. Nonetheless, most of the established risk factors of RSVH have been confirmed in several non-Nordic populations,^24,26^ supporting the generalizability of the model. The availability, effectiveness in clinical use and price of the immunoprophylaxis will also affect how our results will best benefit their targeting. Presently, the model predicts the probability of RSVH for an individual infant, and as such it could be used in e.g. neonatal unit before discharge for every infant, or for estimating the overall RSVH risk in specific subpopulations. The results can also be adapted into even more simple decision rules. The optimal passive immunisation strategy could leverage epidemic timing and intensity predictions combined with real-time epidemiological surveillance data.^38^ Finally, a population-based study after Covid-19 showed that the incidence of severe RSV disease increased in children >1 year,^9^ and if this trend continues after broader RSV immunisation, we should be ready to search for risk factors for RSVH also in older children.

### Meaning of the study

The identified risk factors and prediction model to assess the infant’s risk for RSVH can be used in targeting passive RSV immunisation. The present results can also be utilised in developing more detailed guidelines and recommendations for RSV immunoprophylaxis strategies, as well as in their health economic evaluations. Our results can be useful also in providing guidance to caregivers of small infants regarding non-pharmacologic means of risk management (such as self-isolation) during an RSV epidemic.

## Supporting information

Tripod checklist

## Data Availability

The code of this project (including the clinical prediction model equation) is available in the FinRegistry GitHub at: https://github.com/dsgelab/rsv.
Access to FinRegistry data can be obtained by submitting a data permit application for individual-level data for the Finnish social and health data permit authority Findata (https://asiointi.findata.fi/). The requests are evaluated on a case-by-case basis. Once approved, the data are sent to a secure computing environment Kapseli and can be accessed within the European Economic Area (EEA) and within countries with an adequacy decision from the European Commission. Data dictionaries for FinRegistry are publicly available on the FinRegistry website (www.finregistry.fi/finnish-registry-data).

## Acknowledgements

We are grateful for the Finnish and Swedish children and their families, whose data made this study possible. We thank the FinRegistry team for creating the data for this project, and we acknowledge CSC – IT Center for Science, Finland, for computational resources.

The icons used in the figure 1 are from The Noun Project, under Creative Commons licence (CC BY 3.0) (https://thenounproject.com/icon/family-1034555/, https://thenounproject.com/icon/pregnancy-2741552/ and https://thenounproject.com/icon/infant-4287/).

## Data sharing statement

The code of this project (including the clinical prediction model equation) is available in the FinRegistry GitHub at: https://github.com/dsgelab/rsv.

Access to FinRegistry data can be obtained by submitting a data permit application for individual-level data for the Finnish social and health data permit authority Findata (https://asiointi.findata.fi/). The requests are evaluated on a case-by-case basis. Once approved, the data are sent to a secure computing environment Kapseli and can be accessed within the European Economic Area (EEA) and within countries with an adequacy decision from the European Commission. Data dictionaries for FinRegistry are publicly available on the FinRegistry website (www.finregistry.fi/finnish-registry-data).

## Footnotes

### Author contributorship statement

PV was the coordinator of the project, responsible for the analyses and writing of the study protocol and first draft of the manuscript. PV, AV, EV and AL preprocessed the FinRegistry data. SK was responsible for the administrative and data management work of the FinRegistry project. SJ and PV planned the analysis methods. PV performed the analyses in the Finnish data. SAR, CL. ECO and AS preprocessed the Swedish data and ran the external validation analyses, and CA supervised the validation analyses in Sweden. EH provided guidance in the classification of congenital heart defects. AP and TH performed the XGboost analysis. AP created the web-based risk calculator. PV, SH, SJ wrote the manuscript, with input from all authors. SH initiated the study and was the clinical lead of the project. SH and AG supervised the study. PV is the guarantor for this work and accepts full responsibility for the conduct of the study, had access to the data, and controlled the decision to publish. The corresponding author (PV) attests that all listed authors meet authorship criteria and that no others meeting the criteria have been omitted.

## Supplementary figures

### 1 Overview of analysis methods

**Supplementary figure 1.**
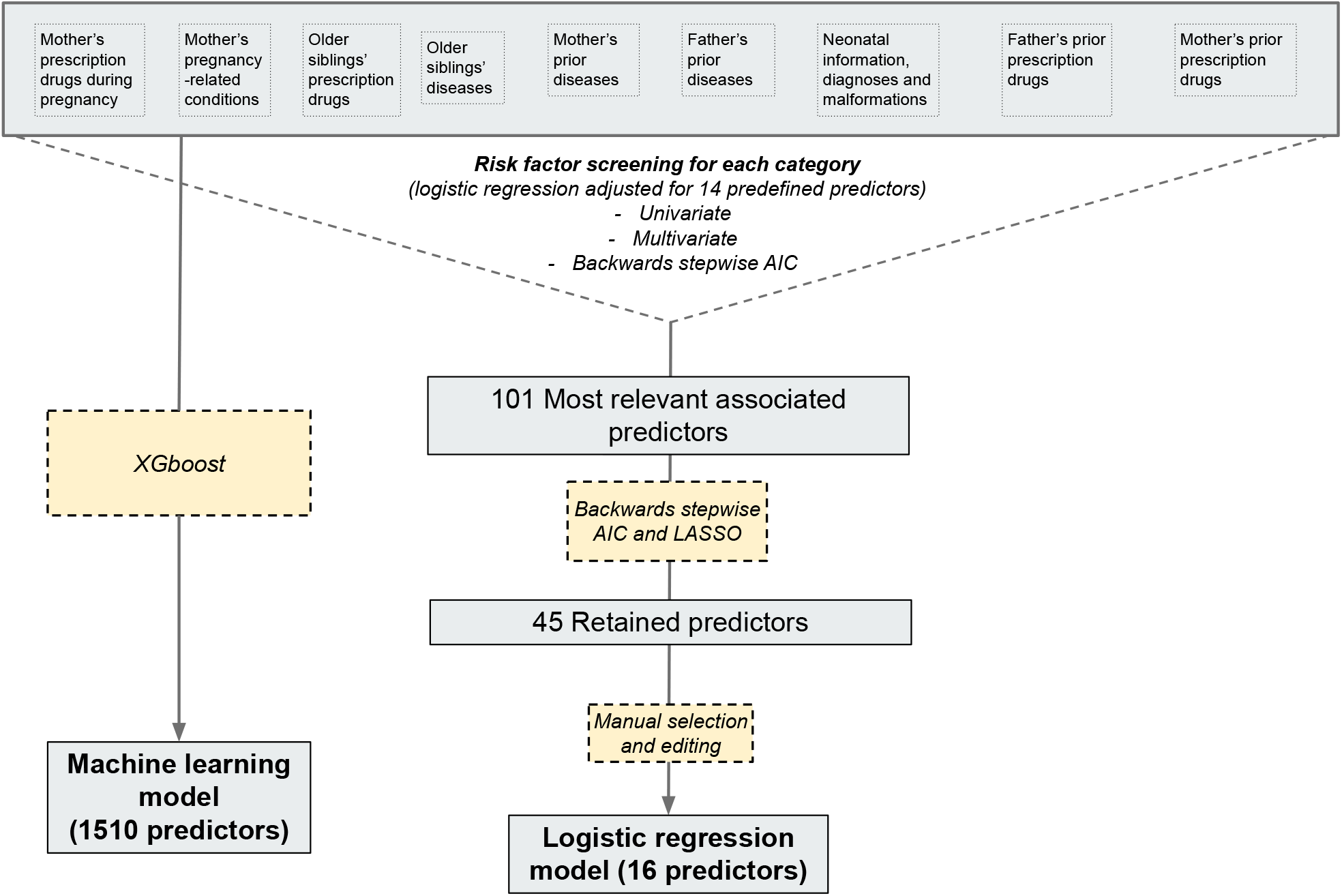
The overview and summary of the analysis methods used in each step of the study. AIC = Akaike information criterion, used as the optimisation metric to guide the backwards stepwise model building. XGboost = eXtreme Gradient boosting, a machine-learning method

### 2 RSV hospitalisations, seasonality and comparison with infectious disease data

**Supplementary figure 2.**
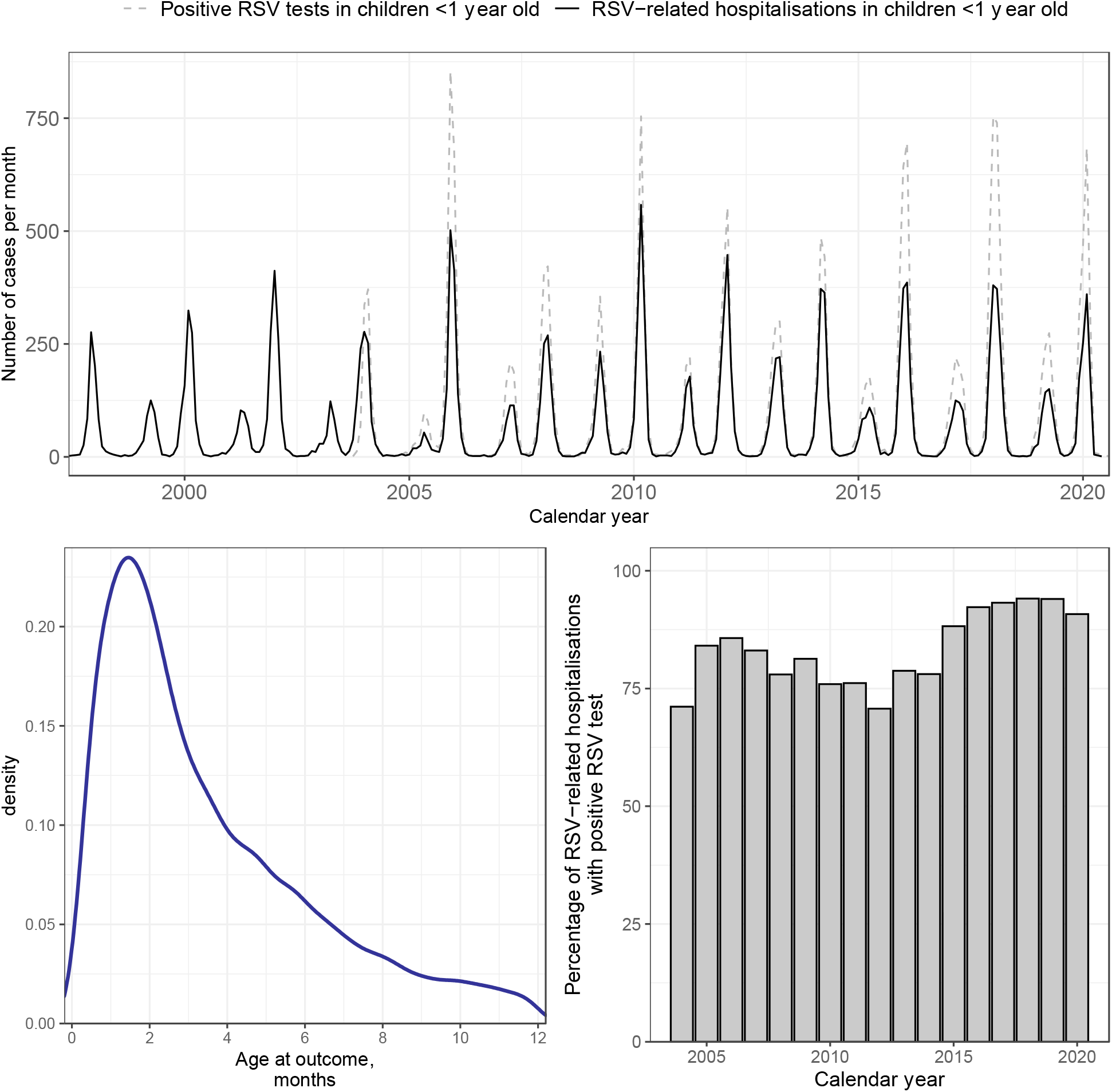
The seasonality (upper panel) and age distribution (lower left panel) of RSV hospitalisations in Finland. In the upper panel, the monthly RSV hospitalisations are compared to the positive RSV test results in children <1 year old in the infectious disease registry. In the lower right panel, the rate of positive RSV test reported to the national infectious diseases register within +/-7 days of RSV hospitalisation is compared during each calendar year. In the lower left panel, the density plot of the age at RSV is shown. RSV hospitalisation occurred typically during the first months of life; Median age at RSV hospitalisation was 80 days (IQR 44-150 days) in Finland and 91 days (IQR 43-224 days) in Sweden.

### 3 Monthly RSV hospitalisation rates in Finland and in Sweden

**Supplementary figure 3.**
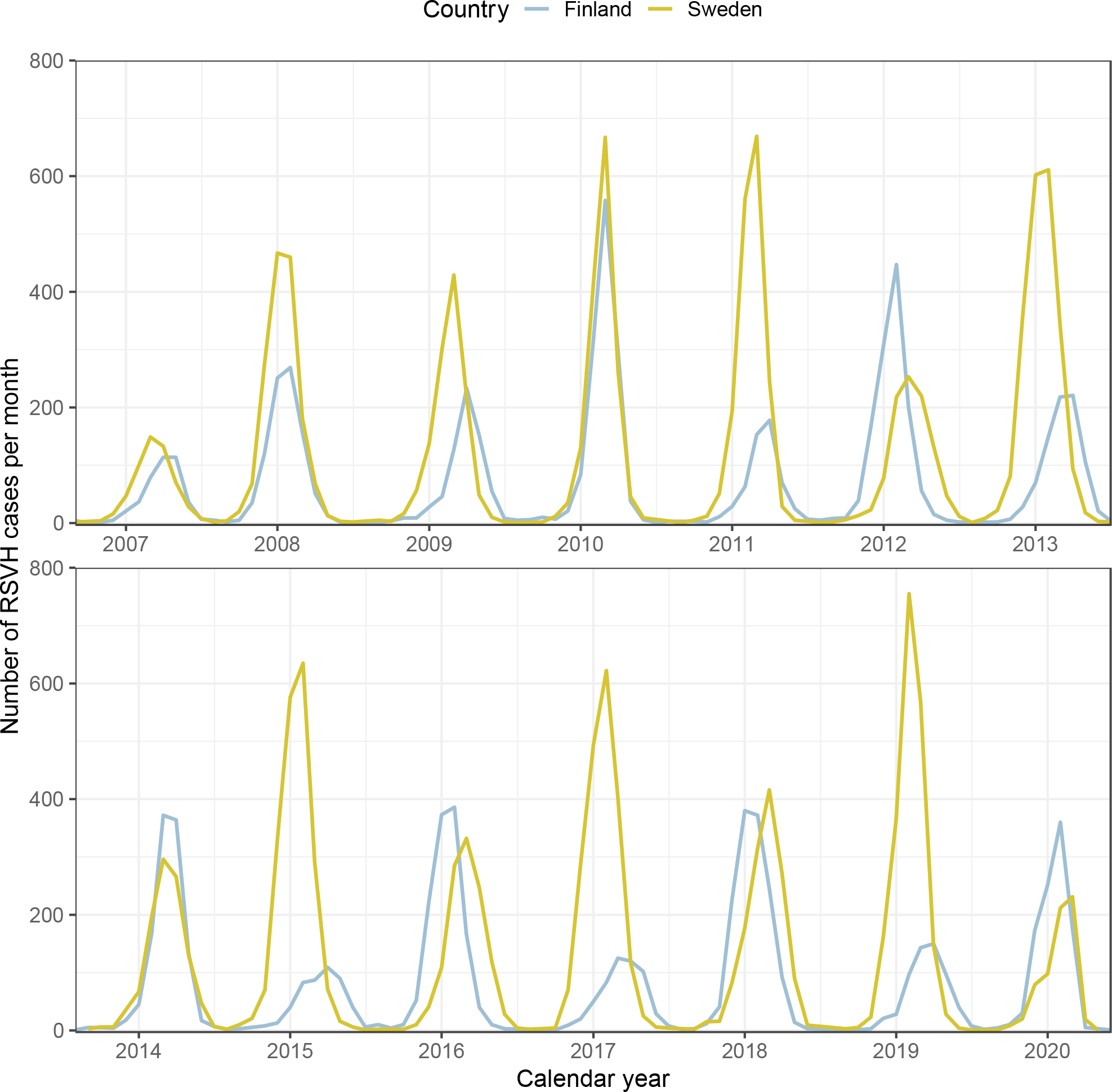
The monthly RSV hospitalisation rates compared between Finland and Sweden. Both countries have some biennial pattern in the epidemic intensity (height of the peaks), but Finland has more distinct biennial variation in the timing of the epidemic peaks. In addition to seasonal variation, we observed a biennial pattern in RSVH, where every other year, the number of hospitalisations peaked earlier and was higher. This biennial pattern diminished towards more recent years. Sweden had similar biennial variation in the epidemic intensity, but the variation in the epidemic timing was smaller. This corresponds to the earlier published reports of RSVH seasonality in the Nordics.

### 4 Histogram of the predicted probabilities

**Supplementary image 4.**
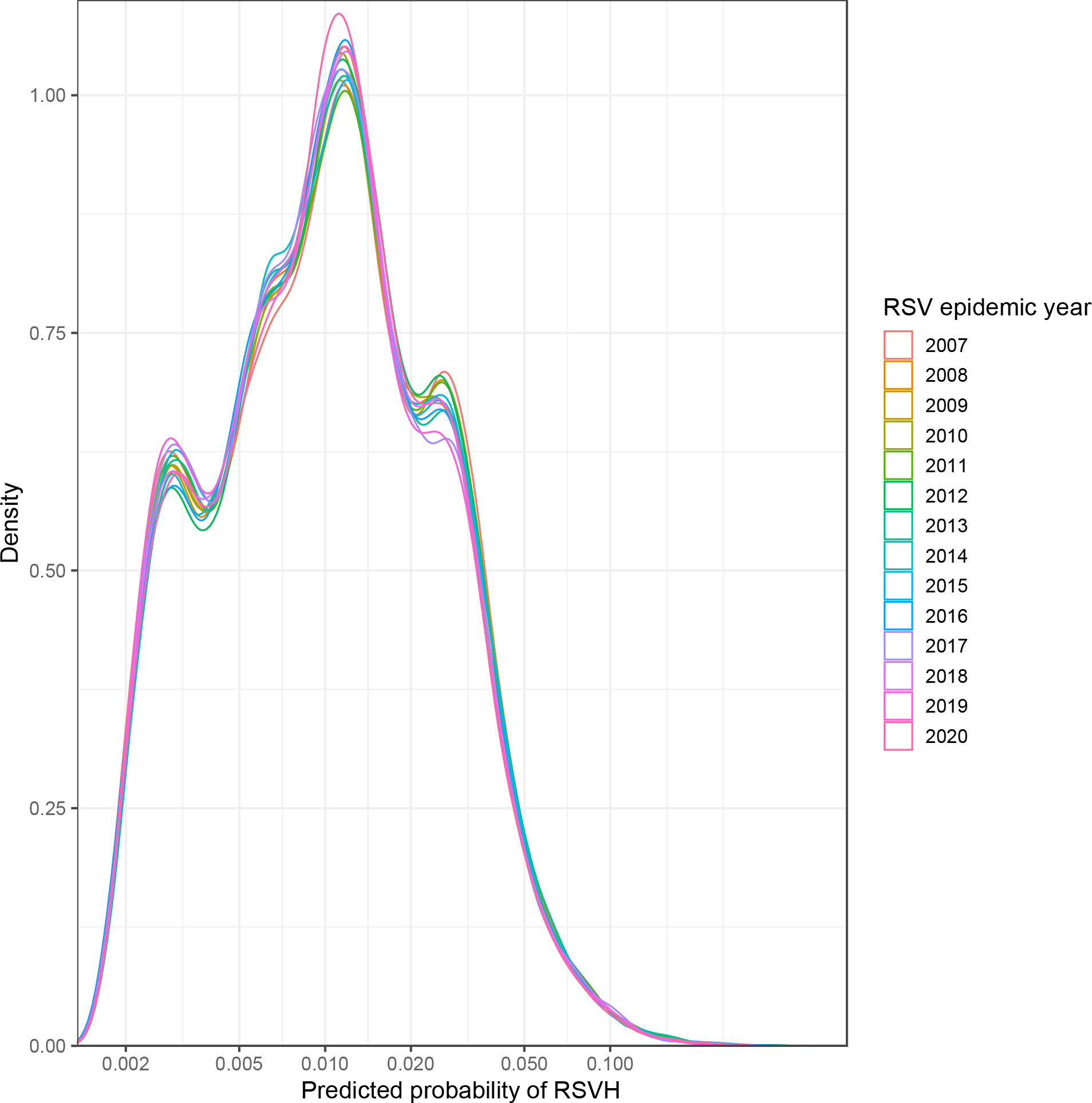
The density plot of the predicted probabilities in the Finnish data, for epidemics 2007-2020. Because of the skewed distribution, the x axis scale is logarithmic. The distribution of probabilities does not significantly vary between years. Supplementary table 15 shows the predicted probability cutoffs for each percentile.

### 5 Population attributable fractions of the model predictors

**Supplementary figure 5.**
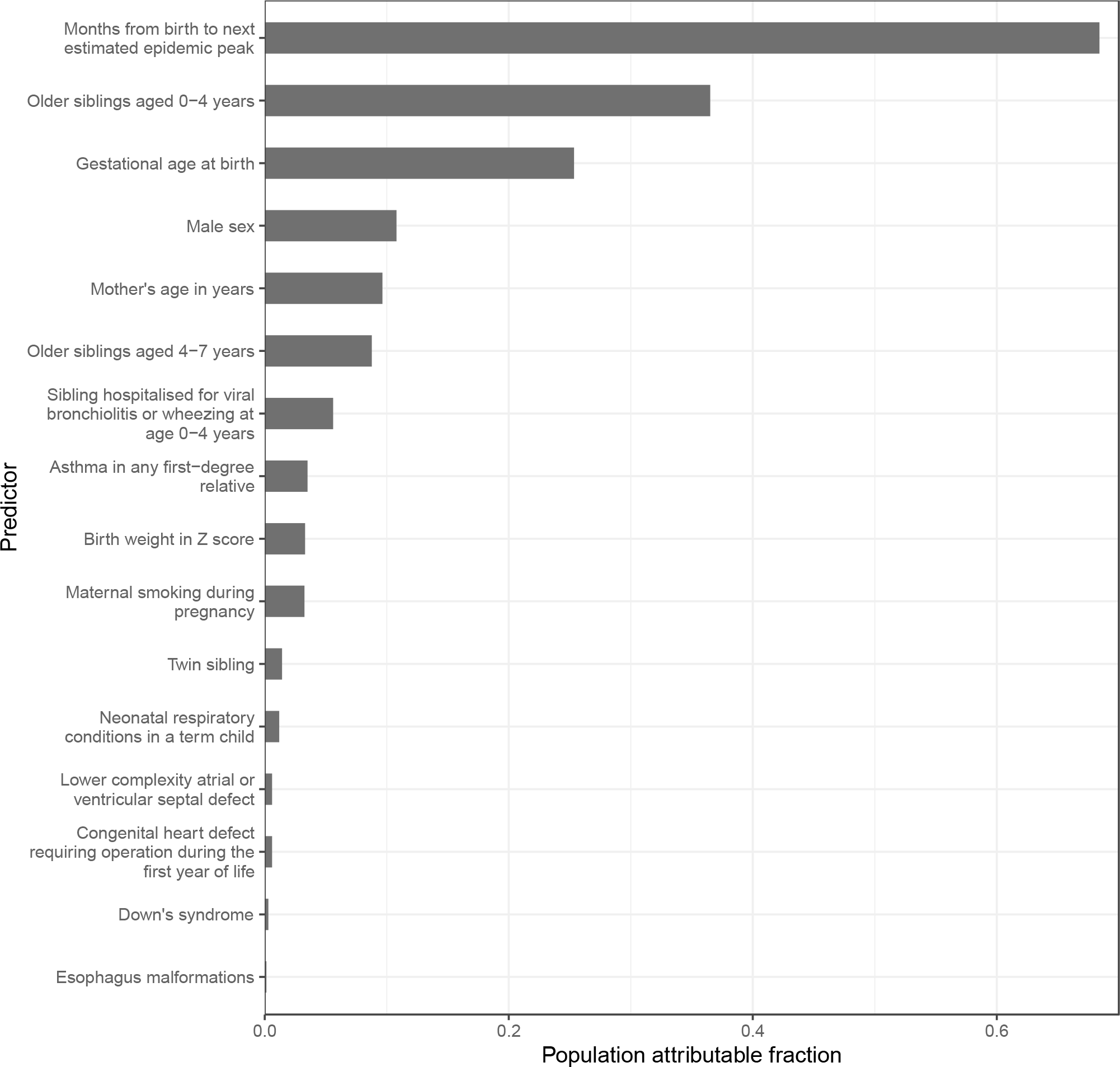
The population attributable fractions for each predictor in the clinical prediction model. The results are obtained from a logistic regression model including the shown variables, and the shown estimates are adjusted for the effect of the other variables. The largest population attributable fractions were observed for months from birth to the next estimated epidemic peak (0.68), having older siblings aged less than 4 years (0.37; aOR 2.42 and 95% CI 2.34 - 2.50) and gestational age at birth (0.25), indicating that these variables were the most impactful at the population-level, reflecting the combination of large effect size and high prevalence of these predictors.

### 6 Discrimination and calibration measures in individual epidemic years

**Supplementary figure 6.**
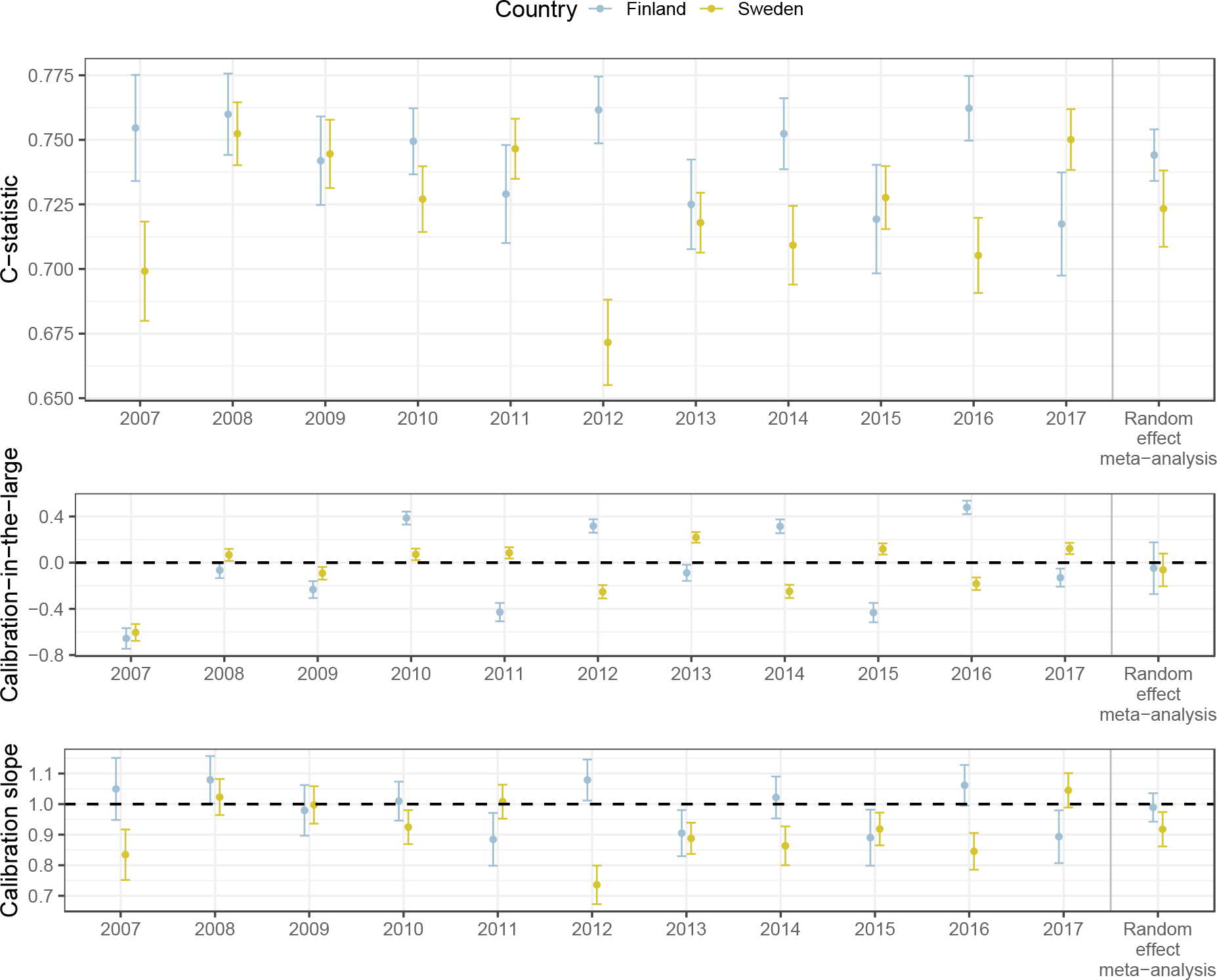
The model performance metrics of the clinical prediction model, shown for the years used in the model training in the Finnish data.

The Finnish metrics are obtained from leave-out cross validation, where each epidemic year is kept as a testing data in turn, the model is trained in all other data and tested in the test fold. The Swedish metrics are from the external test data, and no model training is done in the Swedish data. Note that the metrics, especially the c-statistics are not directly comparable, as in the Swedish data, the timing of the RSV epidemic is estimated from the previous years, and in the Finnish data, the actual epidemic timing is used.

The year in the x axis indicates the RSV epidemics; We used the 1st of June as a start for the RSV epidemic year (for example, all children born between the 1st of June 2007-31st of May 2008 were grouped for RSV epidemic year 2018). In calibration metrics, dashed lines indicate perfect calibration. Random-effect meta-analysis is the meta-analysed metric from all shown epidemic years.

Calibration-in-the-large in the complete pooled external validation data was -0.06 (95% CI - 0.05 to -0.08), and the calibration slope was 0.94 (95% CI 0.92 - 0.95), likely explained by the slightly lower outcome rate in Sweden.

### 7 Discrimination and calibration according to the outcome prevalence

**Supplementary figure 7.**
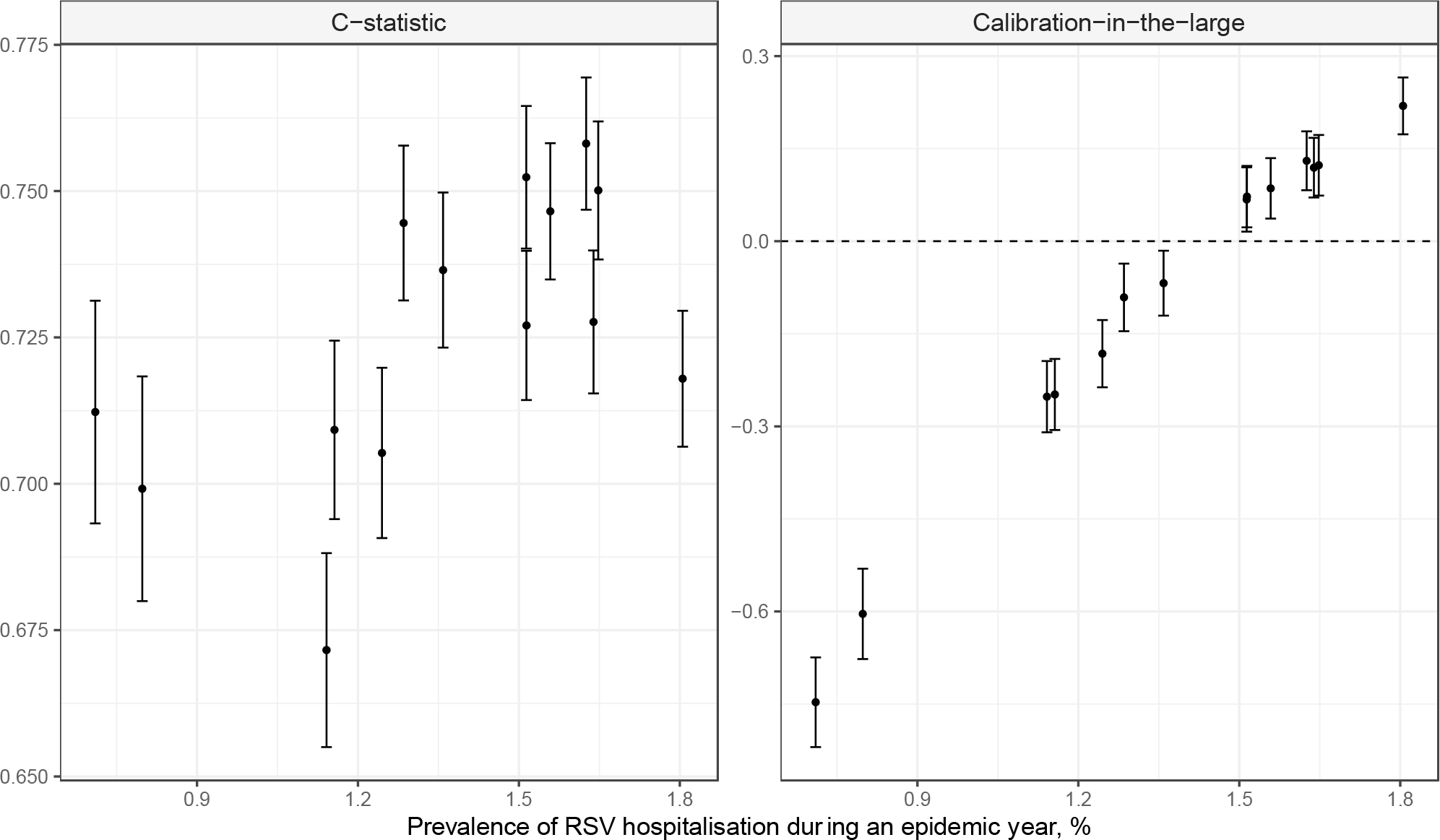
The model performance metrics (C-statistic for discrimination and calibration-in-the-large for calibration) shown according to the prevalence of the outcome, i.e. the percentage of infants hospitalised in Sweden during RSV epidemics 2007-2020. Each dot represents an RSV epidemic year, i.e. children born between June-May. X-axis shows the percentage of the children having the outcome, i.e. severe RSV-LRTI requiring hospitalisation. Y-axis shows the respective performance metric. Dashed line in right panel is the reference for perfect calibration-in-the-large.

### 8 The calibration plots

**Supplementary figure 8.**
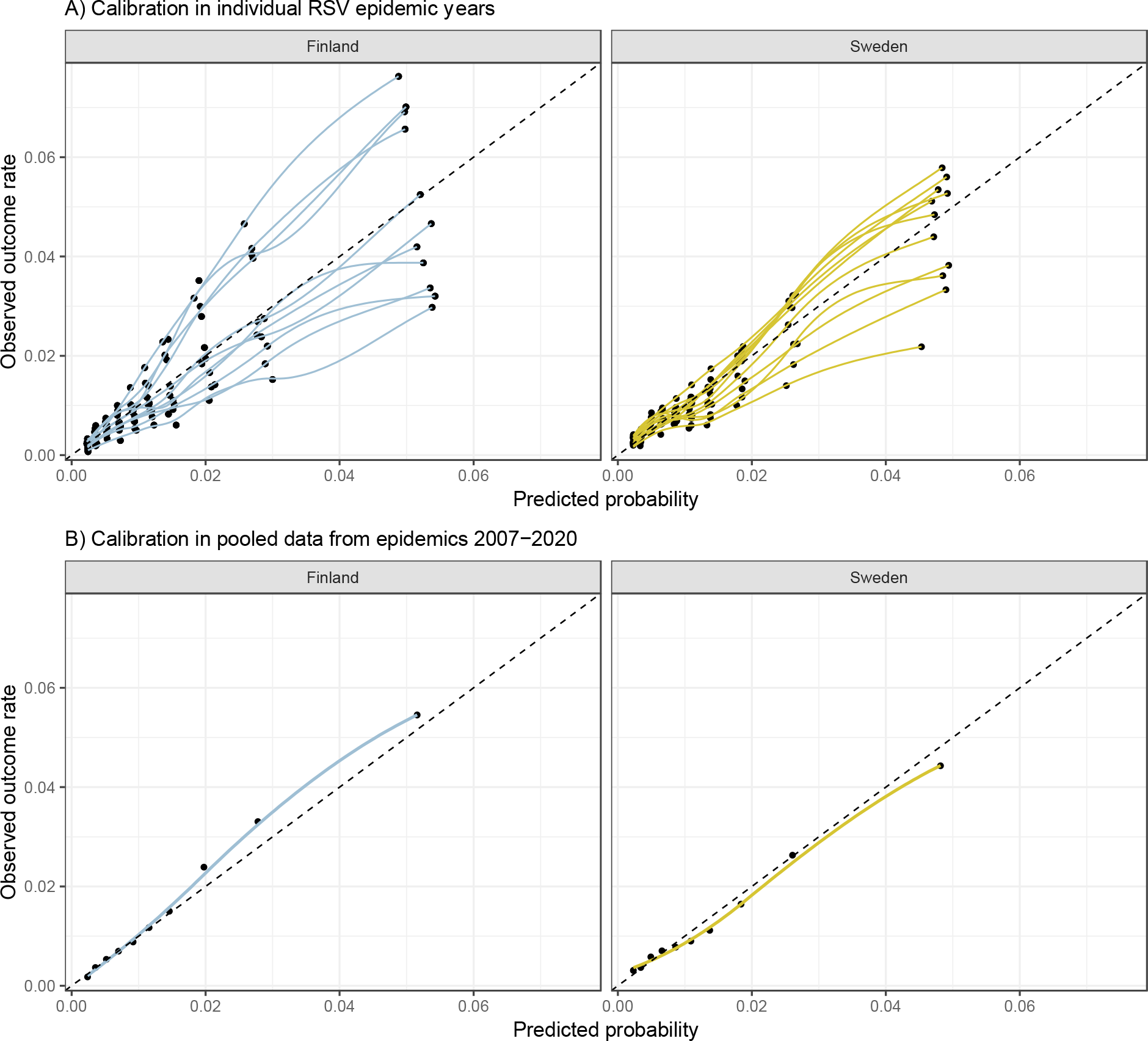
The calibration plots, i.e. the predicted vs the observed probabilities in equally sized deciles divided according to the predicted probability, separately for the three epidemic years of this comparison. Each dot represents a decile, and axes show respectively their mean predicted probability and the mean observed outcome rate

In Panel A, The data for RSV epidemics 2007-2017, corresponding to the Finnish training data, are shown individually for each year. We used the 1st of June as a start for the RSV epidemic year (for example, all children born between the 1st of June 2007-31st of May 2008 were grouped for RSV epidemic year 2018). The dashed lines indicate perfect calibration.The Finnish results are obtained from leave-out cross validation, where each epidemic year is kept as a testing data in turn, the model is trained in all other data and tested in the test fold. The Swedish metrics are from the external validation data, and no model training is done in the Swedish data.

In Panel B, calibration data is shown for pooled data covering epidemic years 2007-2020 in both countries. To obtain these pooled results in Finland, we combined the development and validation data and used the final prediction model to assign predicted probabilities to each individual. Similarly to panel A, the Swedish results are from external validation data.

### 9 Comparison of XGboost and clinical prediction model

**Supplementary image 9.**
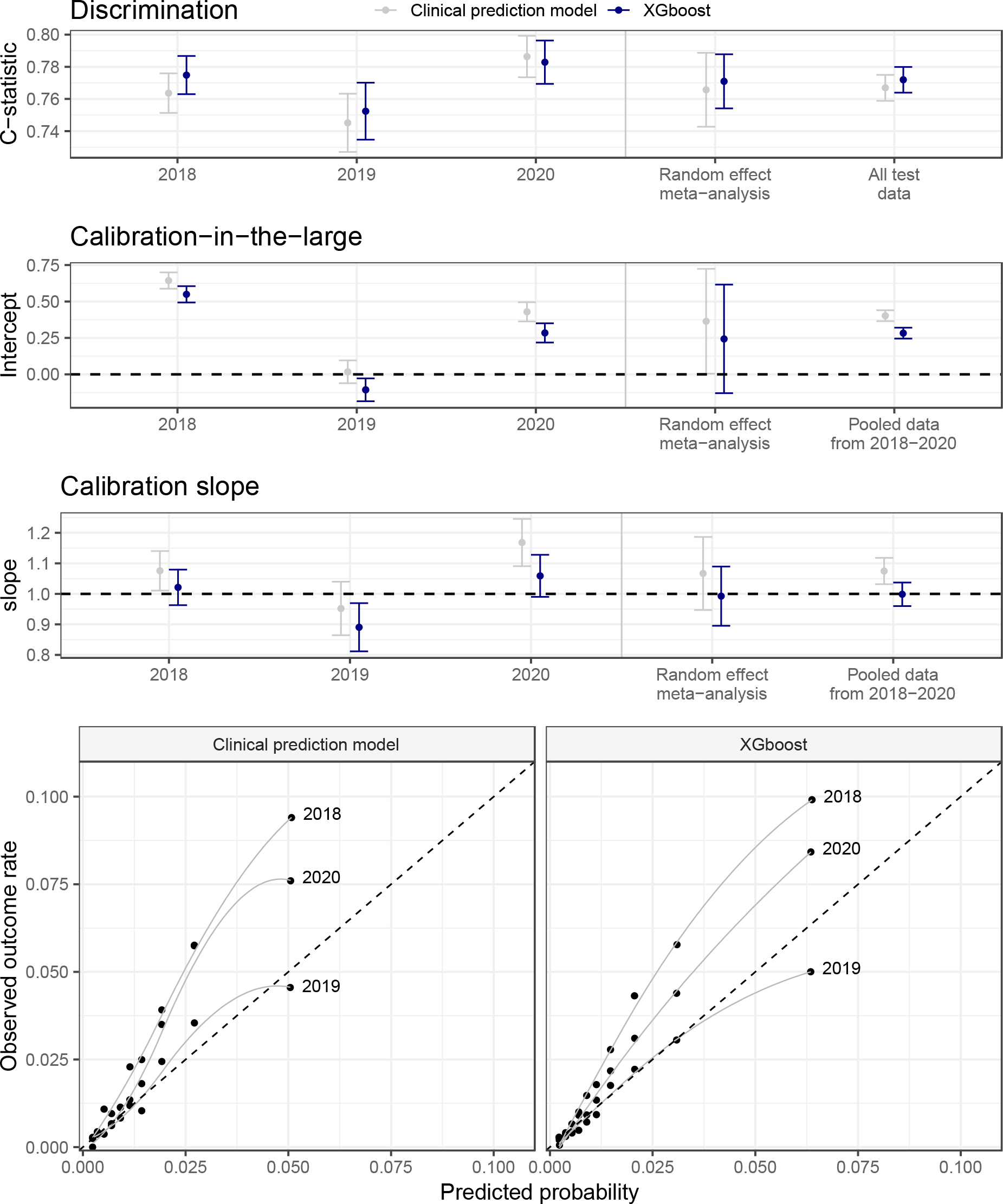
The discrimination and calibration of the XGboost model shown separately for children predisposed to individual RSV epidemic. The year numbers in the image indicate the RSV epidemics. We used the 1st of June as a start for the RSV epidemic year (for example, all children born between the 1st of June 2018-31st of May 2019 were categorised for RSV epidemic year 2019). To summarise the results of the uppermost 3 panels, the respective metrics from each RSV epidemic are combined with random effect meta-analysis. Also the results obtained from testing the model in the complete held-out test set (all 3 epidemics pooled) are shown for comparison. The 2 lowest panels show the calibration curve, with the test data divided into 10 bins and plotting those bins’ mean predicted probability and mean hospitalisation rate against each other. Reference lines show perfect calibration. Note the biennial pattern in the measures, too low predicted probabilities occurring during strong epidemics and lower predicted probabilities in milder epidemics.

### 10 SHAP values of XGboost model

**Supplementary image 10.**
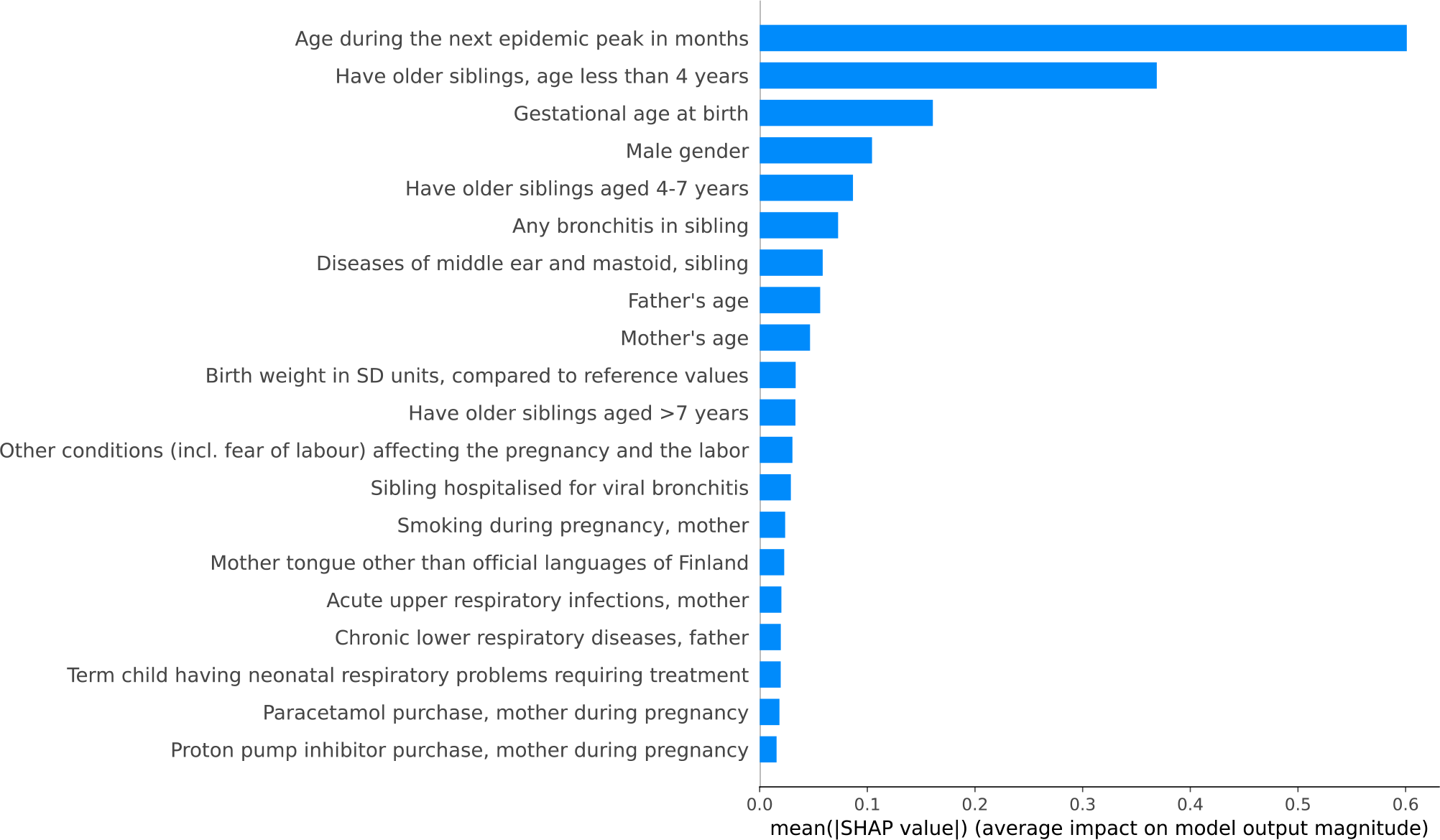
Variable importance in the XGBoost model for the 20 most important variables, based on the SHapley Additive exPlanation (SHAP) values. Higher SHAP value indicates higher predictor importance for the XGboost model.

### 11 Fairness analysis

**Supplementary figure 11.**
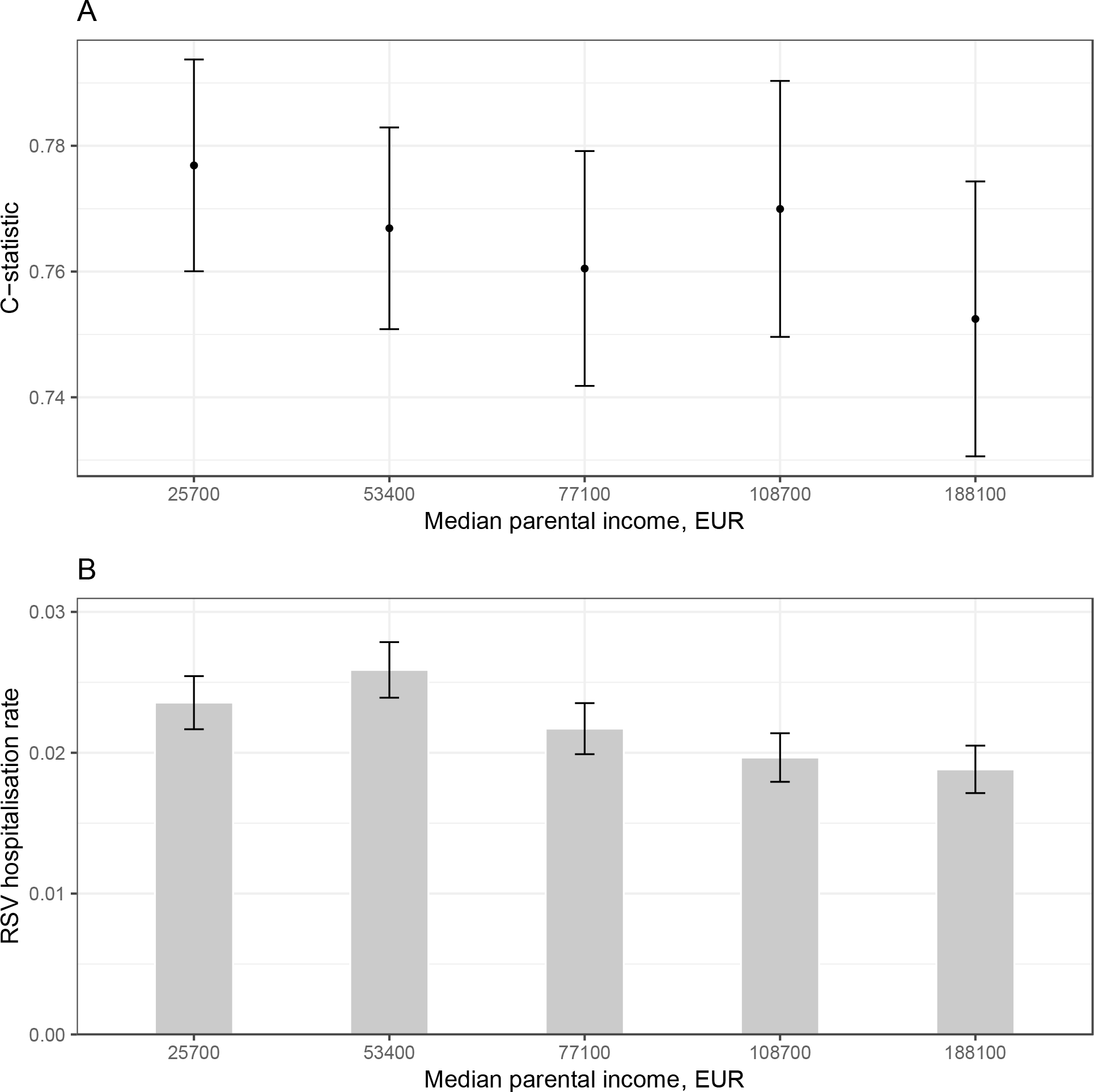
The fairness analysis. Panel a) shows the comparison of the C-statistic between parental income quintiles. For descriptive purposes, also the RSV hospitalisation rates for each quintile are shown in panel b). X axis labels show the median income in each quintile and the error bars show 95% confidence intervals of the c-statistic and RSV hospitalisation rate. The results are from Finnish held-out validation data, covering children born between June 2018 and 5/2020. 5962 children (4.8%) In the Finnish internal validation set had missing parental income. In this population, the C-statistic was 0.746 (95% CI 0.710 - 0.781), slightly worse than in all of the quintiles. The C-statistic in the lowest quintile was 0.777 (95% CI 0.760-0.794), compared to 0.753 (0.731-0.775) in the highest quintile (supplementary figure 11) suggesting that the model performed slightly better in, and thus does not harmfully discriminate against children from lower income families.

## References

1 Li Y, Wang X, Blau DM, et al. Global, regional, and national disease burden estimates of acute lower respiratory infections due to respiratory syncytial virus in children younger than 5 years in 2019: a systematic analysis. The Lancet 2022; 399: 2047–64.

2 Wildenbeest JG, Zuurbier RP, Korsten K, et al. Respiratory Syncytial Virus Consortium in Europe (RESCEU) Birth Cohort Study: Defining the Burden of Infant Respiratory Syncytial Virus Disease in Europe. J Infect Dis 2020; 222: S606–12.

3 Hall CB, Weinberg GA, Blumkin AK, et al. Respiratory Syncytial Virus-Associated Hospitalizations Among Children Less Than 24 Months of Age. PEDIATRICS 2013; 132: e341–8.

4 Broberg EK, Waris M, Johansen K, Snacken R, Penttinen P, European Influenza Surveillance Network. Seasonality and geographical spread of respiratory syncytial virus epidemics in 15 European countries, 2010 to 2016. Eurosurveillance 2018; 23. DOI:10.2807/1560-7917.ES.2018.23.5.17-00284.

5 Renko M, Tapiainen T. Change in respiratory syncytial virus seasonality in Finland. Acta Paediatr 2020; 109: 202–3.

6 Hamrin J, Bennet R, Berner J, Rotzén-Östlund M, Eriksson M. Rates and risk factors of severe respiratory syncytial virus infection in 2008-2016 compared with 1986-1998. Acta Paediatr 2021; 110: 963–9.

7 Obando-Pacheco P, Justicia-Grande AJ, Rivero-Calle I, et al. Respiratory Syncytial Virus Seasonality: A Global Overview. J Infect Dis 2018; 217: 1356–64.

8 Lenglart L, Ouldali N, Honeyford K, et al. Respective role of non-pharmaceutical interventions on bronchiolitis outbreaks, an interrupted time series analysis based on a multinational surveillance system. Eur Respir J 2022; : 2201172.

9 Nygaard U, Hartling UB, Nielsen J, et al. Hospital admissions and need for mechanical ventilation in children with respiratory syncytial virus before and during the COVID-19 pandemic: a Danish nationwide cohort study. Lancet Child Adolesc Health 2023; : S2352464222003716.

10 Mazur NI, Terstappen J, Baral R, et al. Respiratory syncytial virus prevention within reach: the vaccine and monoclonal antibody landscape. Lancet Infect Dis 2022; : S1473309922002912.

11 Hammitt LL, Dagan R, Yuan Y, et al. Nirsevimab for Prevention of RSV in Healthy Late-Preterm and Term Infants. N Engl J Med 2022; 386: 837–46.

12 Madhi SA, Polack FP, Piedra PA, et al. Respiratory Syncytial Virus Vaccination during Pregnancy and Effects in Infants. N Engl J Med 2020; 383: 426–39.

13 Cromer D, van Hoek AJ, Newall AT, Pollard AJ, Jit M. Burden of paediatric respiratory syncytial virus disease and potential effect of different immunisation strategies: a modelling and cost-effectiveness analysis for England. Lancet Public Health 2017; 2: e367–74.

14 Li Y, Hodgson D, Wang X, Atkins KE, Feikin DR, Nair H. Respiratory syncytial virus seasonality and prevention strategy planning for passive immunisation of infants in low-income and middle-income countries: a modelling study. Lancet Infect Dis 2021; 21: 1303–12.

15 Chaw PS, Hua L, Cunningham S, et al. Respiratory Syncytial Virus-Associated Acute Lower Respiratory Infections in Children With Bronchopulmonary Dysplasia: Systematic Review and Meta-Analysis. J Infect Dis 2020; 222: S620–7.

16 Chaw PS, Wong SWL, Cunningham S, et al. Acute Lower Respiratory Infections Associated With Respiratory Syncytial Virus in Children With Underlying Congenital Heart Disease: Systematic Review and Meta-analysis. J Infect Dis 2020; 222: S613–9.

17 Shi T, Balsells E, Wastnedge E, et al. Risk factors for respiratory syncytial virus associated with acute lower respiratory infection in children under five years: Systematic review and meta–analysis. J Glob Health 2015; 5: 020416.

18 Haerskjold A, Kristensen K, Kamper-Jørgensen M, Nybo Andersen A-M Ravn H, Graff Stensballe L. Risk Factors for Hospitalization for Respiratory Syncytial Virus Infection: A Population-based Cohort Study of Danish Children. Pediatr Infect Dis J 2016; 35: 61–5.

19 Heinonen S, Süvari L, Gissler M, Pitkänen O, Andersson S, Helve O. Transient Tachypnea of the Newborn Is Associated With an Increased Risk of Hospitalization Due to Respiratory Syncytial Virus Bronchiolitis. Pediatr Infect Dis J 2019; 38: 419–21.

20 American Academy of Pediatrics Committee on Infectious Diseases. Updated Guidance for Palivizumab Prophylaxis Among Infants and Young Children at Increased Risk of Hospitalization for Respiratory Syncytial Virus Infection. Pediatrics 2014; 134: e620–38.

21 Reeves RM, van Wijhe M, Lehtonen T, et al. A Systematic Review of European Clinical Practice Guidelines for Respiratory Syncytial Virus Prophylaxis. J Infect Dis 2022; 226: S110–6.

22 Manzoni P, Figueras-Aloy J, Simões Eaf, et al. Defining the Incidence and Associated Morbidity and Mortality of Severe Respiratory Syncytial Virus Infection Among Children with Chronic Diseases. Infect Dis Ther 2017; 6: 383–411.

23 Chan M, Park JJ, Shi T, Martinón–Torres F, Bont L, Nair H. The burden of respiratory syncytial virus (RSV) associated acute lower respiratory infections in children with Down syndrome: A systematic review and meta–analysis. J Glob Health 2017; 7: 020413.

24 Jacoby P, Glass K, Moore HC. Characterizing the risk of respiratory syncytial virus in infants with older siblings: a population-based birth cohort study. Epidemiol Infect 2017; 145: 266–71.

25 Figueras-Aloy J, Manzoni P, Paes B, et al. Defining the Risk and Associated Morbidity and Mortality of Severe Respiratory Syncytial Virus Infection Among Preterm Infants Without Chronic Lung Disease or Congenital Heart Disease. Infect Dis Ther 2016; 5: 417– 52.

26 Green CA, Yeates D, Goldacre A, et al. Admission to hospital for bronchiolitis in England: trends over five decades, geographical variation and association with perinatal characteristics and subsequent asthma. Arch Dis Child 2016; 101: 140–6.

27 Collins GS, Reitsma JB, Altman DG, Moons K. Transparent reporting of a multivariable prediction model for individual prognosis or diagnosis (TRIPOD): the TRIPOD Statement. BMC Med 2015; 13: 1.

28 Ferguson J, Maturo F, Yusuf S, O’Donnell M. Population attributable fractions for continuously distributed exposures. Epidemiol Methods 2020; 9. DOI:10.1515/em-2019-0037.

29 Chen T, Guestrin C. XGBoost: A Scalable Tree Boosting System. In: Proceedings of the 22nd ACM SIGKDD International Conference on Knowledge Discovery and Data Mining. San Francisco California USA: ACM, 2016: 785–94.

30 Vickers AJ, Van Calster B, Steyerberg EW. Net benefit approaches to the evaluation of prediction models, molecular markers, and diagnostic tests. BMJ 2016; : i6.

31 Simões Eaf, Madhi SA, Muller WJ, et al. Efficacy of nirsevimab against respiratory syncytial virus lower respiratory tract infections in preterm and term infants, and pharmacokinetic extrapolation to infants with congenital heart disease and chronic lung disease: a pooled analysis of randomised controlled trials. Lancet Child Adolesc Health 2023; : S2352464222003212.

32 Vartiainen, Pekka. Risk of hospitalization from RSV bronchiolitis in a nationwide integrated registry data. 2021. DOI:10.17605/OSF.IO/SWYNH.

33 Straňák Z, Saliba E, Kosma P, et al. Predictors of RSV LRTI Hospitalization in Infants Born at 33 to 35 Weeks Gestational Age: A Large Multinational Study (PONI). PLOS ONE 2016; 11: e0157446.

34 Lejeune S, Sfeir R, Rousseau V, et al. Esophageal Atresia and Respiratory Morbidity. Pediatrics 2021; 148: e2020049778.

35 Nurminen P, Koivusalo A, Hukkinen M, Pakarinen M. Pneumonia after Repair of Esophageal Atresia–Incidence and Main Risk Factors. Eur J Pediatr Surg 2019; 29: 504– 9.

36 Fitzpatrick T, McNally JD, Stukel TA, et al. Family and Child Risk Factors for Early-Life RSV Illness. Pediatrics 2021; 147: e2020029090.

37 Stensballe LG, Kristensen K, Simoes EAF, et al. Atopic Disposition, Wheezing, and Subsequent Respiratory Syncytial Virus Hospitalization in Danish Children Younger Than 18 Months: A Nested Case-Control Study. Pediatrics 2006; 118: e1360–8.

38 Jenkins DA, Martin GP, Sperrin M, et al. Continual updating and monitoring of clinical prediction models: time for dynamic prediction systems? Diagn Progn Res 2021; 5: 1.

