## Supplementary material for "Risk factors for severe respiratory syncytial virus infection during the first year of life: development and validation of a clinical prediction model"

#### Table of Contents

*BACKGROUND* ... 2

**Previous prediction models of severe RSV disease** ... 2

*METHODS* ... 3

**Study subjects and registry data** ... 3

Model development and validation data ... 4

Missing data ... 4

**Sample size calculation** ... 5

**Variable definitions** ... 6

The outcome (RSVH) ... 6

Family member definitions ... 6

Candidate predictor groups and prevalence ... 7

Family members' clinical endpoints ... 7

Pregnancy-related diagnoses, neonatal diagnoses and malformations ... 8

Family members' prescription drugs ... 11

**Statistical analysis methods** ... 12

Restricted cubic spline encoding of continuous variables in logistic regression models ... 12

Interactions ... 12

Lasso ... 12

XGboost ... 13

**Predictor selection** ... 13

Predictor selection inside predictor category ... 14

Multivariable selection ... 14

Variable exclusion criteria ... 14

**Final prediction model training** ... 16

Population attributable fractions ... 16

**Model performance assessment** ... 17

Development and validation data ... 17

Model performance measures ... 17

Decision curve analysis ... 18

Hypothetical immunoprophylaxis targeting ... 18

Fairness ... 18

### BACKGROUND

#### Previous prediction models of severe RSV disease

Several clinical prediction models for severe RSV disease have been developed, but they have considerable limitations.

A model by Rietveld et al.[1] was developed in a Dutch registry-based data of 140 000 children. They used 5 predefined predictors, and trained the model to predict the monthly risk of RSV hospitalisation. The predictors were gestational age, birth weight, sex of the child, bronchopulmonary dysplasia (BPD) and age during the month of prediction. The model predictions were, however, strongly dependent on the local epidemic pattern in the Netherlands, and the model lacks validation in external data; the model was only validated in the training data with re-sampling methods. It is likely that this model will not generalise well to countries with even slightly different epidemic patterns, due to the fixed coefficients of the baseline monthly risk included in the model.

In another study from Netherlands,[2] Houben et al. developed a prediction model from a prospective birth cohort and implemented it as a scoring tool. Predictors were day care attendance and/or having older siblings, parental education level, birth between September-April and birth weight >4kg. However, despite interesting setting, the study had low sample size (298), and lacked validation.

Several prediction models have been developed for preterm infants, and they do not likely generalise to term infants.[3–6]

### METHODS

#### Study subjects and registry data

The study leverages nationwide integrated registry data from the FinRegistry project (www.finregistry.fi). The FinRegistry covers approximately 7,2 million Finnish persons and integrates their registry data regarding health, sociodemographic factors and family relations. This dataset includes 7,166,416 individuals of whom 5,339,804 (74.5%) are index individuals (every resident of Finland alive on January 1st 2010). The remaining 1,826,612 individuals are relatives (offspring, parents, siblings) and spouses of index individuals who are not index individuals themselves.

In this study, the most important of registries were: 1) the Medical Birth Register containing data on newborn babies, their diagnoses after birth and pregnancy-and mother-related variables; 2) Care Register for Health Care (CRHC) of The Finnish Institute for Health and Welfare (THL) which contains data on hospitalizations and contacts to secondary healthcare with diagnoses; 3) Drug purchase register of Kela (the Social Insurance Institution of Finland), containing all prescription drug purchases; 4) population registry of Digital and Population Data services Agency of Finland containing family relations; 5) the Register of Congenital malformations of THL; and 6) the Finnish National Infectious Diseases Register (NIDR) of THL containing data on microbiological samples. The detailed descriptions of FinRegistry and included registries can be accessed at https://www.finregistry.fi/finnish-registry-data.

In FinRegistry, the majority of health-related data are structured into predefined clinically meaningful composite endpoints based mainly on the ICD-classified diagnoses, drug purchases and reimbursements, hospitalisations, causes of death and procedural codes.[7] These endpoint data were used for screening the family members' health conditions.

The Swedish data originated from a register linkage of several national health and population registers including the National Patient Register containing data on ICD-10 codes for all hospital visits and most visits to specialist outpatient clinics, which was used to define the comorbidities. Data on birth weight, gestational age and maternal smoking during pregnancy were retrieved from the Medical Birth Register. Data on asthma medications, which were used for the definition of asthma in the study subjects and their family members, were retrieved from the Prescribed Drug Register.

#### Model development and validation data

The Finnish study population was divided into a development dataset (birth between June 1997 and May 2017,  n = 1 126 952) and an temporal hold-out validation dataset (birth between June 2017 and May 2020, n = 130 352). The full development dataset was used for predictor screening, but the last 10 years were used to estimate the final model coefficients (birth between June 2006 and May 2017, n = 621 487) to maximise generalisation to future years. We assessed the performance of the prediction model in the Finnish hold-out temporal validation set consisting of epidemics 2018-2020 (children's birth date between June 1, 2017 and May 31, 2020). These epidemic years were not considered when training the model or selecting the predictors.

External validation was performed in a nationwide dataset from Sweden in children consisting of epidemics 2007-2020 (children's birth date between June 1 2007 and May 31 2020, n = 1 459 472). The main results are shown in the epidemics 2018-2020, to facilitate comparison with the Finnish data and to illustrate the model performance in the most recent available data.

#### Missing data

Missing values in the predictors were rare. In the Finnish data, 2 602 (0.21%) infants were excluded because of missing information on gestational age or birth weight. Respectively, 205 children (0.16%) were excluded from the Finnish internal validation set because of missingness in either gestational age or birth weight. In the Swedish data, 78 400 (5.1%) had missingness in either gestational age, birth weight or maternal smoking during pregnancy. The missingness was more prevalent in the Swedish data, but as it was used for testing the model only, we refrained from imputation.

Additionally, 39 596 (3.1%) infants were missing their father's ID, and they were not included in the data-driven screening analyses, but were included in the prediction model development and validation (as no directly father-related variables were selected to the prediction model).

The following table details the missingness in each variable.

| | Finland | Sweden |
|---|---|---|
| Sample size (cases with missingness included) | 1 257 515 | 1 537 872 |
| Missingness in clinical prediction model variables, n (%) | | |
| The outcome (RSV hospitalisation) | 0 | 0 |
| Gestational age | 2602 (0.21%) | 304 (0.01 %) |
| Birth weight | 2602 (0.21%) | 1 902 (0.12%) |

| | | |
|---|---|---|
| Mother's age at birth | 0 | 0 |
| Male gender | 0 | 0 |
| Twin sibling | 0 | 0 |
| Older siblings aged 0-4 years | 0 | 0 |
| Older siblings aged 4-7 years | 0 | 0 |
| Down's syndrome | 0 | 0 |
| Sibling hospitalised for viral bronchitis at age 0-4 years | 0 | 0 |
| Mother has smoked during pregnancy | 0 | 76 922 (5.00%) |
| Term child having neonatal respiratory conditions | 0 | 0 |
| Esophagus malformations | 0 | 0 |
| Asthma in a first-degree relative | 0 | 0 |
| Congenital heart defect requiring operation during the first year of life | 0 | 0 |
| Lower complexity atrial or ventricular septal defect | 0 | 0 |
| Missingness in other variables not used in the model, n (%) | | |
| Father's ID | 39596 (3.1%) | (Not analysed) |

#### Sample size calculation

Before starting the data analysis, we did a preliminary sample size calculation for the number of outcomes required to assess a certain number of predictors [8]. In the sample size calculation, we used following assumptions: a) acceptable difference of 0.05 in apparent & adjusted R-squared b) margin of error of 0.05 in estimation of intercept and c) outcome prevalence of 0.016 in events per predictor parameter (EPP) d) desired AUC of 0.75. The estimated EPP was 10.35, and the required sample size for studying 1500 parameters in the development dataset was 970742. However, instead of actually considering the full number of 1500 variables, most variables were excluded from the final model because of their definition or characteristics (see below).

### Variable definitions

#### The outcome (RSVH)

The outcome was defined as hospitalisation with the ICD-10 diagnosis J21.0 meaning RSV bronchiolitis between the age of 7-365 days. Excluding the age 0-7 days from outcome was deemed reasonable from the prediction interval's perspective. Even if prophylaxis was given during the very first days of life, the passive immunisation is unlikely to provide full protection during the first week of life. The rationale for the 1-year follow-up was that severe RSV bronchiolitis develops most commonly during the first months of life, and after the first year of life the risk of hospitalisation is significantly lower. Consequently, the main target population for novel immunoprophylaxis methods are infants entering their first RSV season.

The outcome was defined from the Care Register for Health Care (CRHC). In Finland, all paediatric inpatient care is given in public hospitals, and the CRHC has full coverage of paediatric hospitalisations. Misclassification is in practice only possible through imperfect use of diagnosis codes (e.g., an infant was hospitalised due to RSV bronchiolitis, but did not have J21.0 code recorded for some reason). As rapid antigen detection (RADT) RSV tests have been in routine use in paediatric hospitals throughout the study period and availability of nucleic acid amplification tests (NAAT) has increased in recent years, most children hospitalised with clinical suspicion of RSV bronchiolitis are routinely tested with microbiological RSV tests.

As a post-hoc analysis of outcome validity, we compared the routinely reported positive RSV tests of in the Finnish National Infectious Diseases Register (NIDR) to those hospitalised with RSV bronchiolitis. All clinical microbiological laboratories automatically report all RSV detections to the NIDR for epidemiologic monitoring purposes. However, the infectious disease registry does not have full coverage during the study period, as during the first years of the study period the majority of positive RSV tests were reported to the registry without a personal identification number, and the results could not be linked to the other registry data. This is why we didn't include positive RSV tests to the outcome definition, but rather used it in the post-hoc validation analysis.

#### Family member definitions

FinRegistry data links family members to each other. We identified the mothers, fathers and siblings' data for the infants being followed for RSV infection. Mother's ID was identified from the Medical Birth Register for all children. Father's ID was identified from the population registries, and was missing for 39596 (3.1%) of children. Sibling's IDs were identified from the population registries for both "full siblings" (sharing both parents) and "half siblings" (sharing only one parent). We further used the mother's ID to connect siblings to each other. The rationale for including also half siblings was that even if two siblings share only 1 parent, it is still considerably likely that they cohabit, or at least are in frequent contact with each other, especially if the two siblings are of similar age. This also makes the sibling variable more easily defined in the clinical prediction context. The presence of an alive twin sibling was identified from the Medical Birth Register. Twin sibling's other information was not analysed in this study.

Using these definitions, it is possible that a child is first an “index infant” being followed for the RSV hospitalisation, and then an older sibling to someone else. However, all of the predictors were defined separately for each scenario.

#### Candidate predictor groups and prevalence

For the data-driven screening, the data was first grouped to categories according to the registry as a source of information: 1) mother’s clinical endpoints, 2) father’s clinical endpoints, 3) siblings’ clinical endpoints, 4) the neonate’s diagnoses and malformations and Medical Birth Register information, 6) pregnancy-related maternal diagnoses from the Medical Birth Register and hospital registries, 7) mother’s, father’s and siblings’ drug purchases before the start of pregnancy, 8) mother’s drug purchases during pregnancy and. From these information sources, the candidate features were preselected according to the prevalence, using cutoffs of n = 1500 cases in the training data for the family member’s variables and n = 200 cases for the neonate’s variables. This prevalence-based preselection was done to minimise the risk of overfitting, and because very rare conditions would probably not be feasible for predicting the risk in the general population.

#### Family members’ clinical endpoints

In FinRegistry data, most healthcare-related registry data (disease diagnoses, prescription drugs, causes of death, operations) are coded into clinically interpretable clinical endpoints. These clinical endpoints describe certain medical conditions, e.g. an acute event such as stroke or the onset of chronic diseases, such as cardiovascular disease. The registries from which the main endpoints are created (Care Register for Healthcare, Register of Primary Healthcare Visits, Causes of Death, Drug Purchases, Drug Reimbursements, and Cancer Register) cover multiple decades of data, requiring harmonisation in order to produce comparable and interpretable endpoints. Currently, 3177 clinical endpoints have been defined in collaboration with clinical working groups with domain-specific knowledge about using diagnostic codes in clinical practice. The endpoint library is structured according to the ICD-10 hierarchy, with minor changes when the ICD-8/9 structure is very different from the ICD-10 structure, or when an endpoint is of specific interest.

The clinical endpoints have originally been defined as part of the FinnGen project [7], www.finngen.fi, a research collaboration involving academic and industry institutions, aiming to collect and analyse genomic and health data from 500,000 Finns (10 % of the population). The clinical endpoints are adapted to FinRegistry,and can be explored interactively via the Risteys web portal (https://risteys.finregistry.fi).

##### *Family member asthma*

Registry data contains multiple different variables and endpoints that are related to asthma, such as acute and long-term diagnoses and drug purchases. Definition of chronic asthma was given careful consideration because it was presumed to be linked with the risk of RSVH.

Parent's asthma was defined as having long-term reimbursement of inhalation medication, which requires that the patient fulfils the diagnostic criteria for asthma and adheres to the medication for 6 months. This reimbursement code can be considered as a strict asthma definition in Finland. Another definition for parents' asthma was regular purchases of inhalation steroids (>=5 purchases in 3 last years before the start of pregnancy) and purchases of bronchodilators (>= 3 purchases in 3 years before pregnancy). For siblings, we required a diagnosis of asthma (J45 group) and >2 purchases of inhalation steroids and >= 2 purchases of bronchodilators during the last year.

In Sweden, the definition of asthma was taken from a validated algorithm based on combinations of inhalers for asthma and asthma diagnoses.[9] In order to fulfil the asthma definition, we required fulfilling medication criteria of two or more dispenses of inhaled corticosteroid, leukotriene receptor antagonists or fixed beta2-agonist and corticosteroid combinations (2 weeks between dispenses in children under 4.5 years) or three or more dispenses of short acting beta2-agonists (within 12 months). For children under 4.5 years we required an asthma diagnosis (J45, J46) in combination with fulfilling medication criteria. In older individuals, either asthma diagnosis or medication criteria were sufficient.

##### *Sibling's hospitalisation for viral bronchiolitis or viral wheeze*

We created a composite variable of sibling's hospitalisation at young age (0-4 years) because of severe viral respiratory tract infection, which would capture the sibling's underlying predisposition. Most common phenotypes for this were determined to be RSV bronchiolitis and viral wheeze. Because it may be difficult to determine whether the hospitalisation occurred because of bronchiolitis or viral wheeze, and as in some countries their definitions are considerably overlapping, we combined these disease entities into a single composite variable. In Finland, we used the ICD-10 group J21, which includes the RSV bronchiolitis (J21.0) as well as the viral wheeze in childhood (J21.90). In Sweden, the code J21.90 does not exist and viral wheeze is usually coded J20.9 or J21, and hence we used different definitions between Finland and Sweden to capture the same phenotype in siblings.

#### Pregnancy-related diagnoses, neonatal diagnoses and malformations

The diagnoses from pregnancy, neonatal period and possible congenital conditions were obtained from the Medical Birth Registry and from hospital registries. In addition, the Medical Birth Registry contains several structural variables we included as predefined or candidate predictors.

The so-called neonatal diagnoses were the P chapter diagnoses from the ICD-10 classification. Congenital malformation diagnoses were the Q chapter diagnoses, and pregnancy-related diagnoses were the O chapter diagnoses. The use of these diagnosis codes is specific to the time of pregnancy, neonatal period and congenital conditions. These neonatal, congenital and pregnancy-specific diagnoses were grouped considering their prevalence and clinical similarity, aiming for a prevalence of over 200 per 1 000 000 infants for each predictor.

##### *Months from birth to the next estimated epidemic peak*

Young age during the RSV infection is one of the most important known risk factors for severe RSV-LRTI. This feature is strongly dependent on the local epidemic patterns. We aimed to include this feature to the model in a robust way so that the model coefficients would not be directly dependent on the national epidemic patterns of RSV, and we aimed to keep the model simple enough for use during routine clinical activity. Age during the epidemic was modelled by calculating the distance of birth month to the next estimated epidemic peak in months. The epidemic peak was defined as the calendar month having the highest number of RSV hospitalisations.

However, in the clinical prediction setting, the upcoming epidemic peak timing is not known. In the model training, we used data from the actual epidemic peaks, based on the number of RSV detections and hospitalisations. For the held-out test set and the external validation dataset from Sweden, we estimated the epidemic peaks from the previous years' RSV epidemiological pattern by averaging the epidemic peaks 2 years, 4 years and 6 years ago. This averaging method was used because of the biennial pattern of RSV epidemics in Finland. This averaging method also probably mimics the situation in clinical decision making, when the epidemic peak is not known and no tools exist for its modelling. When using the clinical prediction model presented in the paper, the user can estimate the next epidemic peak with a method and data that is most appropriate in their specific context and location. Importantly, when evaluating model performance in hold-out validation data, we have only used information available at 7 days after birth for each infant.

Months to the next estimated epidemic peak was estimated as the time difference in months between the birth month and the estimated peak month of RSV hospitalisations. We used the actual timing of the peak month in the model training data, but in the validation data, the next epidemic peak was estimated by taking the average peak month from epidemics 2 years, 4 years and 6 years prior to birth in order to account for the biennial variation observed in both countries[10,11], and to restrict the validation analyses to information available at the time of birth of the child.

##### *Birth weight relative to the reference values*

Birth weight is a fundamental recorded attribute of newborn children, and also relevant for RSVH risk. As children are born at different times of pregnancy (some are preterm, some term). It is common practice to record the birth weight in units of standard deviation (SD), i.e. compare the absolute weight to the reference values considering e.g. gestational age at birth.

In Finland, we used the national reference values defined from Finnish population distribution.[12] These reference values consider the gender, gestational age at birth and possible twinhood (twins usually weight slightly less). In Sweden, the Nordic reference values were used.[13] Both reference values are in routine use in recording the birth weights in Finland and Sweden.

##### *Maternal smoking during pregnancy*

Maternal smoking during pregnancy was defined by either having a Medical Birth Register entry indicating any smoking during pregnancy, or the neonate having a diagnosis P04.2 (The effect of mother's smoking on the neonate). In Finland, the maternal smoking variable Medical Birth Register variable contains mention of any smoking history during pregnancy, even during the first trimester, and the information is obtained during routine primary-care based pregnancy checkups or during labour.

In Sweden, maternal smoking during pregnancy was defined as smoking during the first antenatal checkup visit, usually in 8-12 weeks of gestation. Those mothers who quit smoking before that visit were not included in this variable.

##### *Congenital diseases of the infant*

For certain severe congenital conditions of the neonates, such as congenital heart defects (CHD) or Down's syndrome, we confirmed the presence of these conditions by using registry data from the first year of life. In these conditions, the condition is often known already before or immediately after birth or at latest during the initial discharge from hospital, but the diagnosis might be confirmed only several months after birth because of e.g. delays in testing or accurate investigations. This confirmation was done before the development of the final prediction model, and the screening was done based on diagnoses associated with the neonatal period. This confirmation before the final model development was done for the following congenital conditions: BPD, CHDs, Down's syndrome and esophagus malformations.

###### Congenital heart diseases

For hemodynamically significant congenital heart defects (hsCHD), we used the presence of corrective cardiac operations during the first year of life as an indicator for hsCHD. We identified those with a procedure code for cardiac procedures during the first year of life (Nordic Classification of Surgical Procedures, NCSP). Supplementary table 10 lists the operation codes considered.

The rationale for this operation-based classification was that hsCHD consist often of multiple different lesions with several distinct ICD-10 codes being used for one child, and the ICD-10 codes do not contain information on the severity of the lesion, differentiating between hsCHD and non-hsCHD based on ICD-10 codes alone, is not feasible. Furthermore, only scarce and inconclusive evidence describes this risk in more detailed subgroups of hsCHD, such as cyanotic or non-cyanotic hsCHDs [14]. In the presence of hsCHD, the likelihood for a corrective operation during the first year of life can in most cases be determined by a paediatric cardiologist at the initial presentation, even before the accurate diagnosis is confirmed. In Finland and Sweden, palivizumab use in infants with hsCHD is conservative, only the infants with the most severe hsCHD receiving the prophylaxis and it is unlikely to confound the present CHD-related results.

In addition to the severe congenital heart diseases, we defined variables for lower complexity atrial septal defect (ASD) and ventricular septal defect (VSD). To create these variables, we identified those with ICD-10 diagnosis codes for ASD *(Q21.11, Q21.18 or Q21.19)*) and VSD (*Q21.0*) in those who: a) did not have corrective cardiac operations during the 1st year of life, and b) had no other CHD diagnosis codes than ASD or VSD, other than the ICD-10 codes of patent ductus arteriosus (*Q25.0*) or patent foramen ovale (*Q21.11*) which often are incidental findings without hemodynamical significance, and do not need interventions. For the final model, we defined lower complexity CHD (lcCHD) as either having lower complexity ASD or lower complexity VSD.

In Sweden, the severe CHD variable was defined on the basis of ICD-10 diagnoses, as the data of Swedish children did not include information on cardiac operations. We identified those ICD-10 codes for CHD that had operation rates in the Finnish data >50% during the first year of life, and defined the hsCHD in Sweden based on the presence of those ICD-10 codes. Supplementary table 11 shows the operation rates observed in Finland for each ICD-10 diagnosis code used to define the hsCHD in Sweden. The prevalence rates using both definitions were similar (**table 1**).

#### Family members' prescription drugs

In the prescription drug purchases, the ATC codes were truncated with the first 5 characters of the ATC code to reflect drug groups. Some 5-digit ATC groups were further grouped if this was clinically justified. For example, the bronchodilator group included the 5-digit ATC code R03AC and the inhalation steroids group included ATC codes R03BA and R03AK.
For parents, we require by default 5 or more purchases during the last 3 years before the child's birth to count that variable as. However, as some drugs are not used regularly but might signify the presence of significant illness, for some drugs we required fewer purchases. For the following drugs, fewer purchases during the 3 last years were required: Injection adrenaline (2 or more purchases); peroral steroids (3 or more); inhaled bronchodilators (3 or more); antihistamines (4 or more); antibiotics (3 or more). For siblings, we required by default 3 or more purchases during the year before the child's birth, and for the aforementioned temporary drugs, 1 or more purchase was required.

### Statistical analysis methods

#### Restricted cubic spline encoding of continuous variables in logistic regression models

In logistic regression analyses, the continuous variables were coded with restricted cubic splines. In the feature selection, we used 4 knots in all continuous variables placed in the default percentiles (5th, 35th, 65th and 95th) [15]. Because the knot placement in quantiles depends on the data used in the *rcs* function, we prespecified and fixed the knot locations for the final model fit in order to produce a model object and equation that could be used in predicting with new and external data.

In the final model, we used 4 fixed knots in default percentiles (5th, 35th, 65th and 95th) for other continuous variables except for the gestational age. Regarding gestational age, the majority of children born term or close to term and the quantile-based knots were automatically based to close to full gestational age, but the most significant risk of severe RSV-LRTI is observed in very preterm infants. This is why we used the 4 default percentile-based knots (placed in 257, 276, 283, 293, gestational days) and 2 manual knots in 203 and 238 gestational days to ensure that the model would adequately capture the risk differences in the more preterm babies. Adding these 2 manual knots to the gestational age lead to the reduction of AIC by 100, calculated in a model containing only spline-transformed continuous variables. Equal AIC reduction of 100 was observed when modelling the gestational age with 6 quantile-based knots.

#### Interactions

We explored the possible interactions by adding individual pairwise interaction terms to the clinical prediction model, and calculating the likelihood ratio test between the model with and without the interaction term. Interaction terms were tested only for the most common variables and where the interaction was clinically plausible. **Supplementary table 9** shows the tested interactions and the respective chi-squared statistic from the likelihood ratio test. The strongest interaction was observed between the age during the next epidemic peak and gestational age, and also other variables had possible interactions with the age during the next epidemic peak. However, as the age during the next epidemic peak is subject to uncertainty and variation across different regions and epidemic patterns, we decided not to include any interaction terms in order to avoid model overfitting. Ultimately, no interaction terms were included in the final model.

#### Lasso

As a part of the predictor selection,, we trained a least absolute shrinkage and selection operator (Lasso) regression model with the potentially relevant candidate predictors identified in the initial feature screening. Lasso (L1-regularisation) was chosen because of its tendency to regularise variable coefficients to 0, i.e., to exclude irrelevant variables. The magnitude of regularised regression coefficients were analysed for the decision to include or exclude variables from the final model. The Lasso model was trained in 10-fold random-split cross validation in the training set, in contrast to the internal validation assessment, done with CV across epidemic years. The lambda value, i.e. the value of the regularisation parameter, was chosen to yield the most regularised model where the cross-validated prediction error was within 1SD of the minimum obtained error. This more conservative value for lambda was chosen to avoid overfitting, and to retain only the most important predictors. In the Lasso model, continuous variables were included as linear terms instead of splines.

##### XGboost

We used an eXtreme Gradient boosting (XGboost, [16]) approach including all 1 510 risk factors to reach the upper limit of predictive information in the data, to use as a benchmark for the clinical prediction model, and also to compare the performance of a complex model including interactions to a simple logistic regression model.

The XGboost model was trained in the 10 most recent epidemics (6/2007-6/2017) of the training data and tested in the Finnish hold-out test set, but the machine learning model could not be validated in the Swedish dataset.

Hyperparameters were optimised with RandomSearchCV and GridSearchCV functions (scikit-learn) over the range of possible hyperparameter values reported in **supplementary table 12**. The optimization was done in the model training dataset using 5-fold cross validation. First, we searched a wider range of hyperparameters with the random search method, and then confirmed the position with the grid search method using narrower range around the values identified by the random search. We did not use class weights and did not consider the classification results of the model, but we analysed only the probabilities of RSV hospitalisation as the model output. SHapley Additive exPlanations (SHAP) values [17] were used to identify the most important predictors in the XGboost model.

#### Predictor selection

First, the association of each candidate predictor was assessed with logistic regression adjusted for the 14 predefined predictors. In summarising the results, statistical significance was set to a $p<4e-5$ after Bonferroni multiple-testing correction because of 1205 tests in the family member variables and $p<1e-3$, because of 255 tests in the child-related variables. Different p value thresholds were used because of the greater number of potentially interesting but rare conditions in the neonate, and because of the exploratory nature of this association analysis. However, the p value was never the only criteria to determine variable inclusion to the prediction model.

When exploring the difference of the predictor effects (i.e. regression coefficients) in mothers and fathers (in figure 1c), we used the Z-test. In this analysis, $p<0.01$ was considered statistically significant due to the exploratory nature of the analysis, and in order to broadly visualise the differences in the regression coefficients between parents.

To select risk factors to include in the final prediction model, we first selected the most relevant variables from each predictor category (infant’s, parents’ and siblings’ diagnoses and drug purchases; see above for details). We then proceeded with multivariate comparisons with the variables selected from each category. These selection processes are described next.

#### Predictor selection inside predictor category

The selection within each category (Mother's, father's, siblings' and the infant's diagnoses and drug purchases; described above) was based on the following three parallel logistic regression results:

1. Univariate odds ratio separately for each variable, adjusted for predefined predictors
2. Multivariate odds ratio, adjusted for the predefined predictors and all other variables from the risk factor category
3. Backwards stepwise elimination of variables from the multivariate model created in the previous step, based on the Akaike Information Criterion (AIC) improvement

#### Multivariable selection

Based on the previous set, we selected the most important candidate predictors for further consideration. The most relevant candidate predictors from each category and the predefined predictors were compared in a multivariable setting. This multivariable comparison was done with two logistic regression models, containing all selected candidate predictors and the predefined predictors.

1. Backwards stepwise elimination of variables based on AIC improvement
2. L1-regularised LASSO model.

From these two models, we assessed the following statistical parameters for each variable: 1) the results of this multivariate comparison (i.e., the multivariate odds ratio and its statistical significance); 2) the possible exclusion from the stepwise model; 3) the regularisation in the LASSO model; 4) the chi-squared statistic from the likelihood ratio test and 5) the prevalence (i.e., rarity) of the predictor.

Supplementary tables 2-7 show the results of the first step (the analyses inside each category, also including all variable definitions and prevalences), and supplementary table 13 shows the results of the multivariate selection results, where most relevant variables from different categories were combined.

#### Variable exclusion criteria

Our aim was to only include features that would be easily defined in the clinical prediction context, that would have similar definitions across time and across different hospitals and countries, that would not have reporting bias due to registry limitations and where a potential mechanism of association was plausible to reflect the probability of severe RSV infection, and not a secondary cause affecting admission decision.

As the most important criterion for predictor selection, we considered whether the predictor's characteristics and definition justify its inclusion to the clinical prediction model. It has been shown that a model based on secondary care data might not perform as well in the primary-care -based population [18]. Furthermore, several registry-based variables might not be suitable for clinical prediction purposes or they might be inconsistently reported depending on the access to secondary or tertiary care.

In the feature selection process, we used case-by-case consideration when deciding the inclusion or exclusion of each variable. We removed predictors where the registry based definition would be difficult to translate clearly to the clinical context, such as family members' antibiotic purchases. We also discarded predictors where the association with the outcome is likely not due to disease severity, but parent or healthcare system related factors (e.g., mother tongue, parent's anxiety disorder diagnoses) that plausibly affect the probability of hospitalisation without necessarily having an effect on disease severity. For the final model, Independent variables were combined into a composite variable if they were related and their odds ratios were similar.

The most important arguments for excluding the variables from the prediction model are listed in the following table.

| | |
|---|---|
| Suspicion that the mechanism of association between the feature and RSV hospitalisation is unclear in nature, reflecting e.g. socioeconomic factors, treatment-seeking behaviour of parents or reduced admission threshold instead of the severity of the RSV-LRTI | Mother's mental-health-related diseases; Mother's drug abuse during pregnancy; family members' income |
| The diagnosis may not capture all with the condition of interest, because of potentially biased reporting in the registries (primary-care -related diagnoses not fully covered in FinRegistry) or because not all individuals with the condition get the diagnosis | Acute otitis media of siblings; Diagnosis of pregnancy-related fatigue |
| Variable having potentially different definitions across time or in different countries. | Socioeconomic factors; Symptomatic or non-specific diagnosis codes; use of certain drugs |
| Vague or unclear variable definition for clinical prediction purpose | Mother's paracetamol purchase; family member depression medication purchase; Antibiotic purchase |
| Overlapping variables - the most clearly defined variable is kept | Chronic lower respiratory conditions vs. any asthma diagnosis vs. long-term drug reimbursement for asthma |
| Statistical criteria | Small odds ratio; rare predictor; small or non-existent Akaike Information Criterion improvement; Regularised coefficient in LASSO model |

For example, one of the most important reasons for excluding a predictor was that the present data do not fully cover primary care records of the patients. The incomplete coverage means that the diagnoses given in primary care are not consistently captured or might have bias in their reporting. For example, sibling's otitis media diagnoses are likely not fully captured in the FinRegistry data, and those cases with the registry entry of otitis media might represent the most severe cases of the disease spectrum. The variable and its coefficient would thus not represent the real-world association with the infant's RSV hospitalisation risk, and the variable was excluded from the prediction model despite significant association. To further exemplify, the diagnosis of upper respiratory tract infection of the family member might be dependent on the treatment-seeking behaviour or access to care of the family.

### Final prediction model training

After selecting the final predictors in the training set, we fitted the logistic regression model in the 10 most recent epidemics of the training set (epidemics between 2007-2017) to estimate the final model coefficients. Continuous variables were included as restricted cubic splines with 4-6 knots. The knots were fixed for prediction in the external data. See above for more in-detail description of the spline method used.

We explored possible interactions by including interaction terms with the strongest risk factors and if the interaction was clinically justified. We assessed each interaction with likelihood ratio test comparing models with and without the interaction term (supplementary table 9).

#### Population attributable fractions

We described the importance of final prediction model variables in relation to each other in the population level with the population attributable fractions (PAF) [19,20]. The PAF describes the fraction of cases in a population attributable to a certain variable, or alternatively, how much the observed cases would have decreased if the effect of this variable were completely removed. We did not aim e.g. to estimate the treatment effect with the PAF calculations, but its purpose was descriptive. The PAF estimates were adjusted for the other variables in the prediction model.

To calculate PAF for the continuous variables, we used the method described by Ferguson et al [20]. The PAF estimates were calculated from a logistic model containing the 16 predictors, continuous variables coded with natural splines with 3 degrees of freedom (instead of restricted cubic splines as in the clinical prediction model), because the R functions required the use of natural spline functions and non-missing data.

### Model performance assessment

#### Development and validation data

The full development set was used in the predictor screening (1 126 952 Finnish infants). After selecting the final model predictors, we obtained the model coefficients from the 10 most recent epidemics of the development set (training data, 621 487 Finnish infants). Finnish hold-out temporal validation data consisted of the infants born between June 2017 - May 2020 (n = 130 352). External validation data consisted of 1 459 472 Swedish children, born between June 2006 and May 2020.

The model performance in the training data (Finnish infants born between June 2006 - May 2017) was tested using so-called leave-one-out cross validation. In this method, one epidemic year at a time is held out for testing, and the model is trained with all remaining data. This process was iterated for each epidemic year of the training data. Thus, in all testing scenarios the model was tested in different data than in which it was trained

In all testing scenarios, we calculated the discrimination and calibration measures in individual epidemic years, and we used a random effects meta-analysis to combine the estimates across the three epidemic years. In parallel with the meta-analysis estimates, we also obtained the discrimination and calibration measures from the pooled testing data for comparison.

#### Model performance measures

Discrimination was assessed with the C-statistics and its 95% confidence interval was obtained from using DeLong method [21]. Calibration plots were created by dividing the predicted probabilities into deciles (10 equal-sized groups) and plotting the observed outcome rate in each of the deciles. Calibration slopes were calculated by training a logistic regression model for the outcome (RSV hospitalisation), with the logit-transformed predicted probability obtained from the prediction model as the only predictor, and examining the linear coefficient as the calibration slope. Calibration-in-the-large, i.e. the overall calibration metric was obtained from a logistic regression model, where the outcome (RSV hospitalisation) was predicted with the prediction model's logit-transformed probability as the offset term (i.e., forcing its coefficient to be 1). The intercept of this regression model is the calibration-in-the-large. 95% confidence intervals for calibration slope and intercept were obtained from standard errors of the regression coefficients.

Discrimination and calibration were assessed in individual epidemic years, to illustrate the variance in model performance in different epidemics. For overall calibration and discrimination, we did random-effect meta-analysis from the metrics of the individual epidemic years, and also calculated the validation metrics in the complete validation dataset.

#### Decision curve analysis

In calculating net benefit, the trade-off between correctly predicting cases (true positives) and incorrectly predicting non-cases as positive (false positives) is weighted according to the threshold probability. The net benefit value reflects this quantified trade-off for certain probability cutoff. The decision curve analysis compares the net benefit across the range of realistic probability cutoffs, and compares different decision making strategies (common references being intervention for all and intervention for none). In the RSV immunisation context the probability range was determined to be 0-15%.

We adapted the American Academy of Pediatrics (AAP) recommendations for palivizumab prophylaxis [22] as a reference prediction strategy. There are no national guidelines for palivizumab use in Finland, although the use of palivizumab in Finland is more conservative than recommended by the AAP guidelines. To reflect these recommendations in our data, we used the following three conditions: gestational age 29 weeks or less; having hemodynamically significant congenital heart defect; and having bronchopulmonary dysplasia. If any of these were fulfilled, the child was defined as fulfilling the AAP criteria for palivizumab.

#### Hypothetical immunoprophylaxis targeting

We assessed the model's potential clinical impact by estimating the effect of prophylaxis on risk of RSVH if the prophylaxis were targeted using predictions from the model. To assess discrimination, we used different percentiles of predicted probability as cutoff for the prophylaxis. We estimated a hypothetical NNT value for each cutoff,separately for each country and each epidemic year of the hold-out validation data to account for the differences in outcome prevalence. We assumed that the efficacy of the immunoprophylaxis was 60% in preventing hospitalisations, chosen as a conservative estimate based on pooled analysis of data from two RCTs including preterm and term infants where the combined efficacy against RSVH was 77.3%.[23–25]

#### Fairness

We analysed the so-called algorithmic fairness, i.e. the possible difference or bias in the performance of the prediction model in vulnerable subgroups, by calculating the C-statistic of the clinical prediction model across parental income quintiles. This analysis was only done in the Finnish internal validation set because the Swedish external test set did not contain income data.

The information on parental income was obtained from the Earnings Register of Finnish Centre for Pensions. We calculated the overall income of the full calendar year before the calendar year of the child's birth. The year prior to the birth of the child was selected in order to reduce the confounding effect of childbirth and the child's possible disease conditions on the parental income. We summed the mother's and father's incomes together to a parental income variable. The incomes were reported in euros. All numbers were corrected for the consumer price index. All these analyses were done in the held-out test set, meaning that we considered income from years 2016-2019. Finally, the incomes were divided into quintiles. Hospitalisation rates with 95% confidence intervals and the median incomes with interquartile ranges were plotted.

In the Finnish internal validation set, 5 962 children (4.8%) have missing income. In this population, the C-statistic was 0.746 (95% CI 0.710 - 0.781), slightly worse than in all of the quintiles.

#### Deviations from the original study protocol

Prior to starting the analyses, we archived a study protocol, available at https://osf.io/h7r9b. The following deviations from the study protocol were made. We updated the study inclusion period to 31.5.2020, because we had data available until the end of 2021. However, as the RSV season in 2021 and 2022 was exceptional because of COVID-19 restrictions, we did not consider including those children born in proximity of this time period to the follow-up. Those born at the end of 5/2021 were followed until May 31st, 2022. The study protocol also doesn't mention the external validation in the Swedish data, as this was confirmed only after the publication of the protocol.

### SUPPLEMENTARY RESULTS

#### Predictor discovery - associations from the predictor screening analyses

We systematically screened for predictors by testing the association between 1496 candidate predictors from parents, siblings and the infant with the infant's RSVH, adjusting for the 14 predefined predictors with logistic regression.

Statistically significant associations were observed for neonatal respiratory conditions, such as transient tachypnea of the newborn (TTN, aOR 1.23, 95% CI 1.12 - 1.34), and "other and unspecified neonatal breathing problems" (aOR 1.41, 95% CI 1.27 - 1.58), but respiratory distress syndrome diagnosis was not associated with increased risk for RSVH (aOR 1.09, 95% CI 0.94 - 1.25) when adjusting for  the 14 predefined predictors (including gestational age at birth). Bony and muscular malformations (ICD-10 codes Q67 and Q68) were associated with an increased RSVH risk (aOR 2.54, 95% CI 1.53 - 5.27). Regarding certain rare conditions considered as risk factors [26], the Q79 diagnosis group including diaphragmatic hernia (aOR 1.09, 95% CI 0.61 - 1.93) and respiratory organ malformations diagnoses Q30-Q34 (aOR 1.11, 95% CI 0.61 - 2.04) were not significantly associated with RSVH.

Among the parents' prior diagnoses, psychiatric diseases and substance use disorder diagnoses, both in mothers and fathers, were clearly associated with increased RSVH risk (**figure 1**). The highest aORs were observed for opioid use disorder diagnosis in fathers

(2.53, 95% CI 2.01 - 3.19) and mothers (2.43, 95% CI 1.78 - 3.22). Substance use of the mother were reflected in the pregnancy-related and neonatal variables through high aORs for diagnoses of withdrawal symptoms of the neonate (2.68, 1.77 - 4.07) and other effects of maternal addictive drug use on the neonate (3.08, 1.98 - 4.79).

Asthma-related variables in all family members, but especially in older siblings, were associated with the risk of RSVH. Sibling's regular montelucast medication, often indicating treatment-resistant or severe allergic asthma, had an aOR of 2.45 (with 95% CI 2.08 - 2.89). Similarly sibling's asthma (aOR 1.75, aOR 1.67 - 1.84), mother's asthma (aOR 1.53, 95% CI 1.41 - 1.65) and father's asthma (aOR 1.22, 95% CI 1.10 - 1.34) were associated with increased risk for RSVH.

Several antibiotic purchases of family members had significant association with RSVH, such as cephalexin during pregnancy (aOR 1.34, 95% CI 1.27 - 1.43). Similarly, common infectious disease diagnoses of the sibling, such as acute otitis media (aOR 1.70, 95% CI 1.63 - 1.77) and gastroenteritis (aOR 1.60, 95% CI 1.50 - 1.69) were associated with the infant's RSVH. Among mother's drug purchases during pregnancy, valproate was most clearly associated with RSVH (aOR 2.45, 95% CI 1.88 - 3.19), probably reflecting the known teratogenic effect of valproate. In fathers, valproate was not significant (aOR 1.18, 95% CI 0.84 - 1.66).

#### Hypothetical number needed to treat (NNT) calculations for immunoprophylaxis targeting

To evaluate the potential utility of the model in different clinical scenarios we estimated the NNT to prevent one RSVH for an immunoprophylaxis targeted according to the prediction model risk percentiles in the validation data from epidemics of 2018-2020 (**Figure 5b)**. We assumed 60% efficacy of the immunoprophylaxis in preventing hospitalisations.[23,24]

In the Finnish validation data, the top 10% infants with highest predicted risk of RSVH (top 90th percentile), the observed RSVH risk was 3.3 times higher (7.3%, vs 2.2%) than in all infants during the 2018-2020 epidemics, and the RSVHs in this top 10% accounted for 33% of all RSVHs. The NNT in this top 10% highest risk group of infants would be 23 (ranging from 18 in 2018 to 36 in 2019), and the prophylaxis would have prevented 20% of all RSVHs (ranging from 18% in 2019 to 21% in 2018). Similarly in Sweden the top 10% highest predicted risk infants had 3.3 times higher observed risk of RSVH than all infants (4.2% vs 1.3%). The RSVHs in these top 10% accounted for 32% of all RSVHs. The NNT would have been 40 (ranging from 29 in 2019 to 77 in 2020), and the prophylaxis would have prevented 19% of all RSVHs (ranging from 18% in 2020 to 20% in 2019).

For comparison, if in Finland the immunoprophylaxis were targeted to the top 1% (the 99th percentile) of children, the NNT estimate would have improved to 14 but only 3% of all hospitalisations would have been prevented. Additionally, assuming the immunisation were targeted according to AAP criteria during 2018-2020 in Finland, the prophylaxis would have been given to 0.6% of infants with NNT of 39 and only 0.6% of all RSVHs would have been prevented.

#### Clinical prediction model equation

The R implementation of the prediction model equation, along with other code of this project, can be found in GitHub, through direct link https://github.com/dsgelab/rsv/blob/main/model_equation.R. The model equation file contains variable names and descriptions in the code comments, in order to facilitate possible future implementation regardless of the platform used.

### SUPPLEMENTARY FIGURES

#### 1 Overview of analysis methods

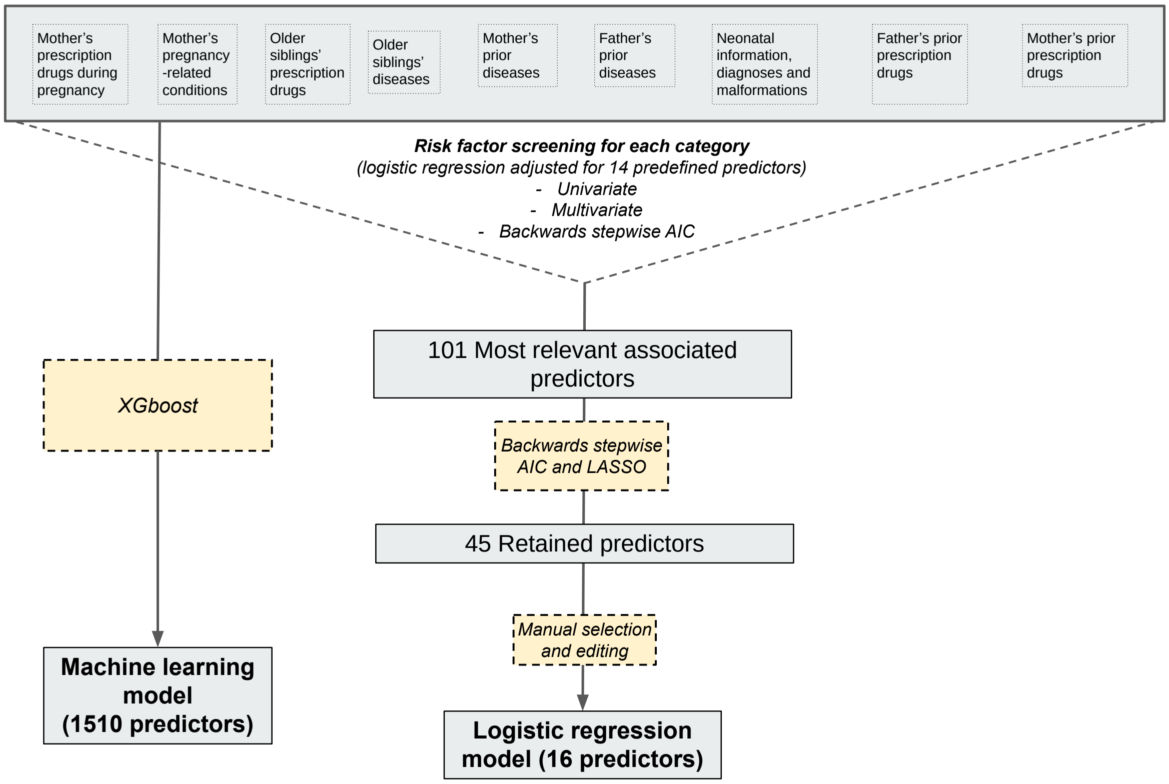

**Supplementary figure 1** The overview and summary of the analysis methods used in each step of the study.
AIC = Akaike information criterion, used as the optimisation metric to guide the backwards stepwise model building.
XGboost = eXtreme Gradient boosting, a machine-learning method

### 2 RSV hospitalisations, seasonality and comparison with infectious disease data

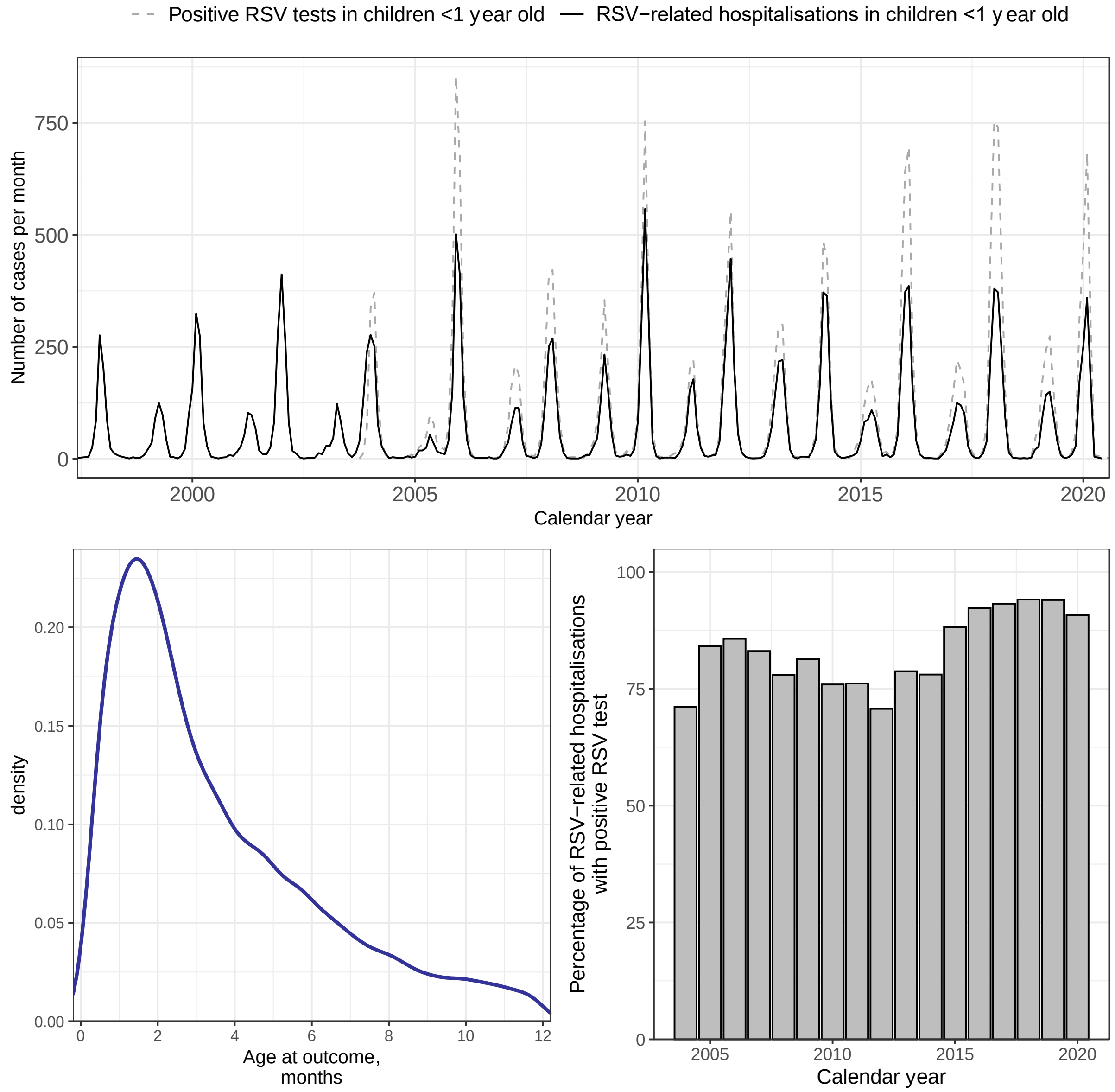

**Supplementary figure 2** The seasonality (upper panel) and age distribution (lower left panel) of RSV hospitalisations in Finland. In the upper panel, the monthly RSV hospitalisations are compared to the positive RSV test results in children <1 year old in the infectious disease registry. In the lower right panel, the rate of positive RSV test reported to the national infectious diseases register within +/- 7 days of RSV hospitalisation is compared during each calendar year. In the lower left panel, the density plot of the age at RSV is shown. RSV hospitalisation occurred typically during the first months of life; Median age at RSV hospitalisation was 80 days (IQR 44-150 days) in Finland and 91 days (IQR 43-224 days) in Sweden.

### 3 Monthly RSV hospitalisation rates in Finland and in Sweden

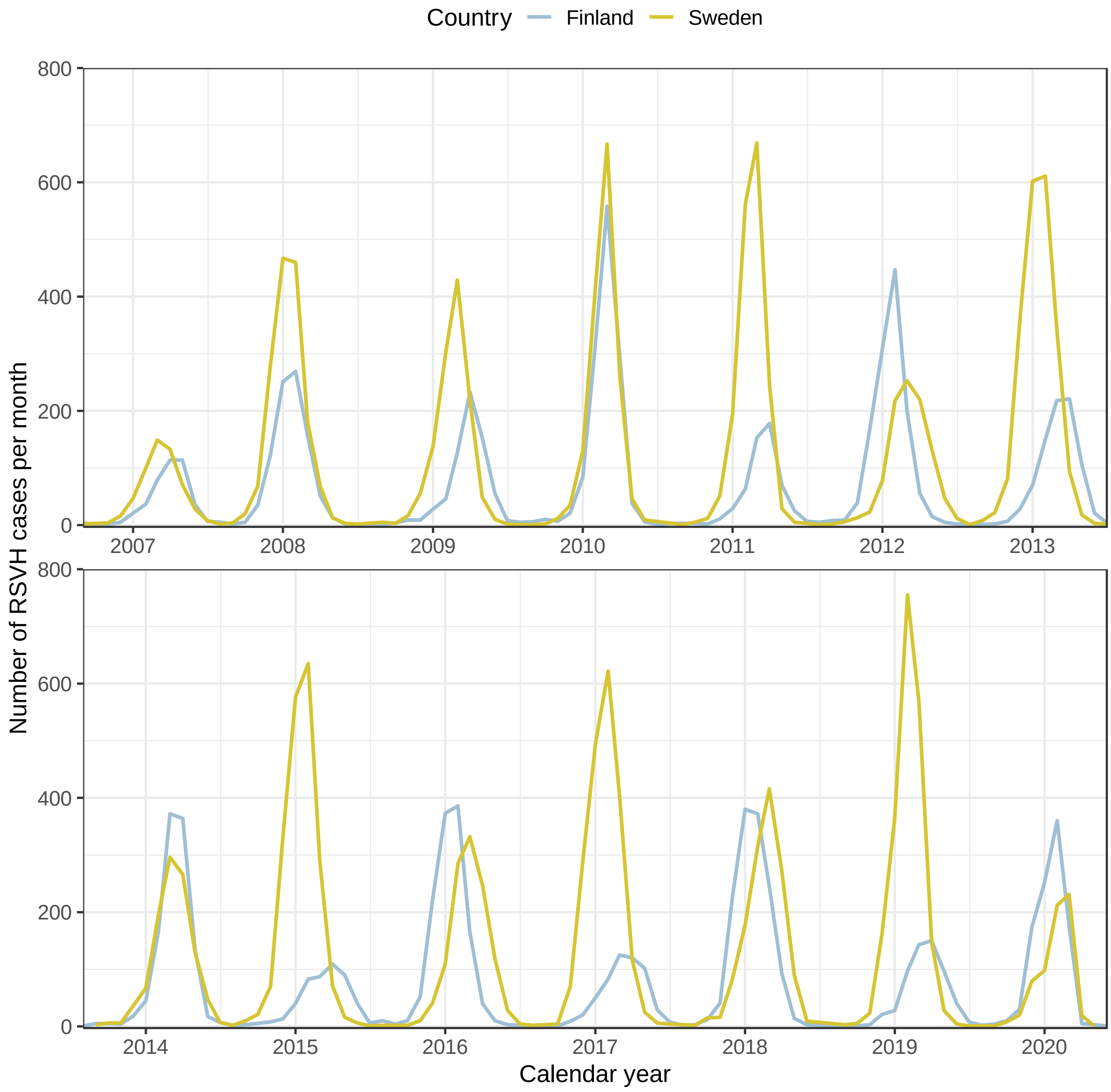

**Supplementary figure 3** The monthly RSV hospitalisation rates compared between Finland and Sweden. Both countries have some biennial pattern in the epidemic intensity (height of the peaks), but Finland has more distinct biennial variation in the timing of the epidemic peaks. In addition to seasonal variation, we observed a biennial pattern in RSVH, where every other year, the number of hospitalisations peaked earlier and was higher. This biennial pattern diminished towards more recent years. Sweden had similar biennial variation in the epidemic intensity, but the variation in the epidemic timing was smaller. This corresponds to the earlier published reports of RSVH seasonality in the nordics [10,11]

### 4 Histogram of the predicted probabilities

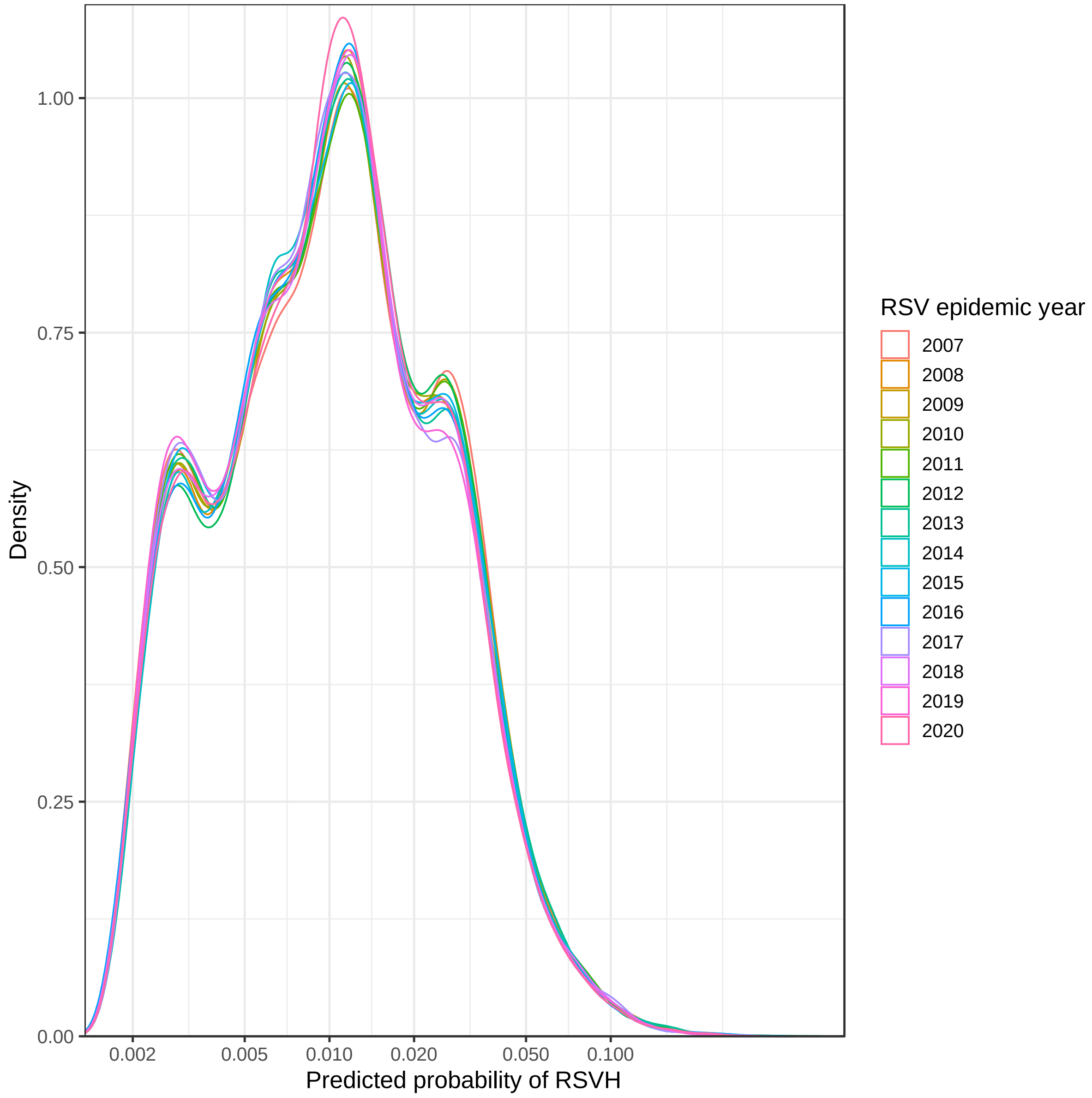

**Supplementary image 4** The density plot of the predicted probabilities in the Finnish data, for epidemics 2007-2020. Because of the skewed distribution, the x axis scale is logarithmic. The distribution of probabilities does not significantly vary between years. Supplementary table 15 shows the predicted probability cutoffs for each percentile.

### 5 Population attributable fractions of the model predictors

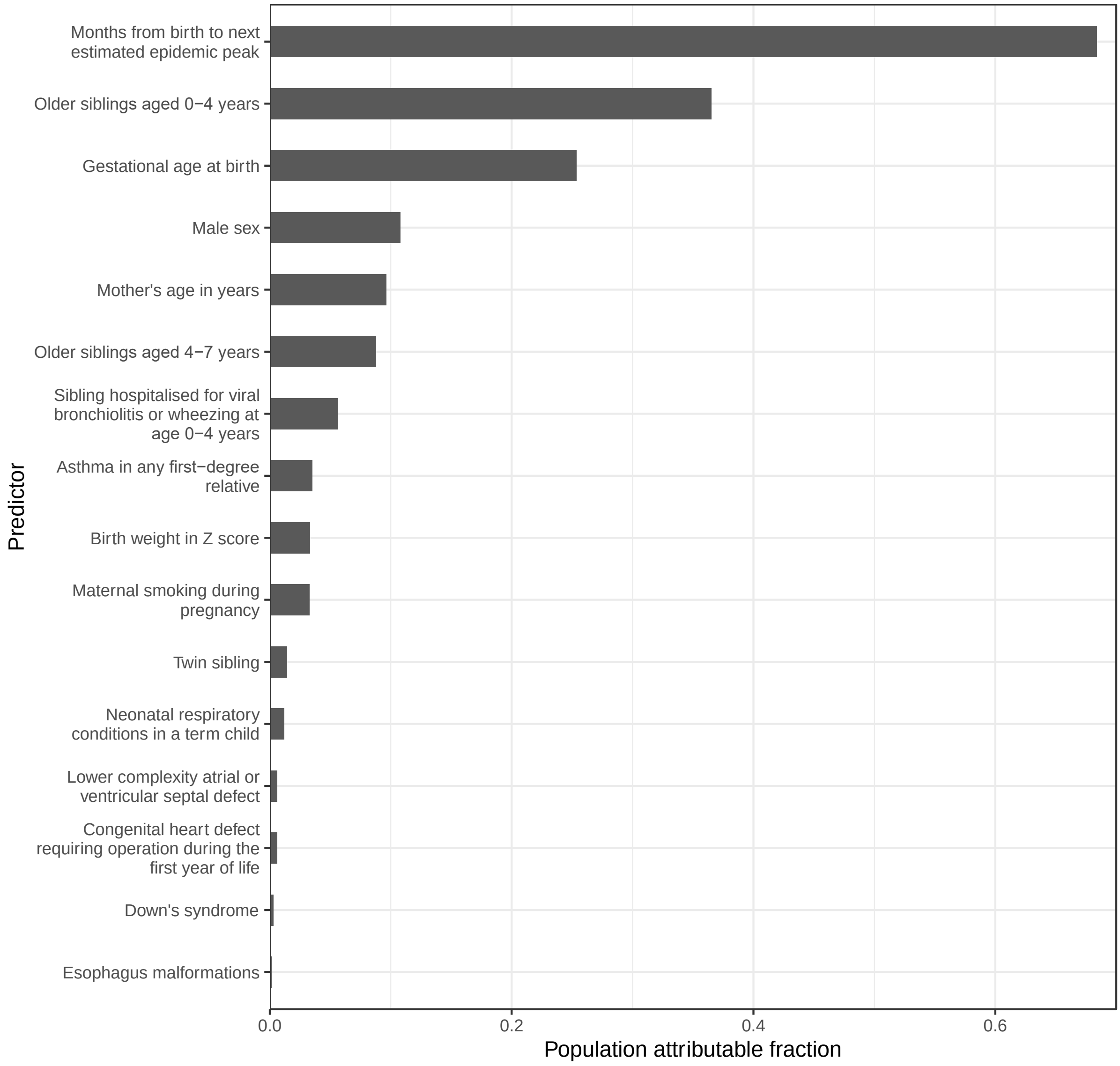

**Supplementary figure 5** The population attributable fractions for each predictor in the clinical prediction model. The results are obtained from a logistic regression model including the shown variables, and the shown estimates are adjusted for the effect of the other variables. The largest population attributable fractions were observed for months from birth to the next estimated epidemic peak (0.68), having older siblings aged less than 4 years (0.37; aOR 2.42 and 95% CI 2.34 - 2.50) and gestational age at birth (0.25), indicating that these variables were the most impactful at the population-level, reflecting the combination of large effect size and high prevalence of these predictors.

### 6 Discrimination and calibration measures in individual epidemic years

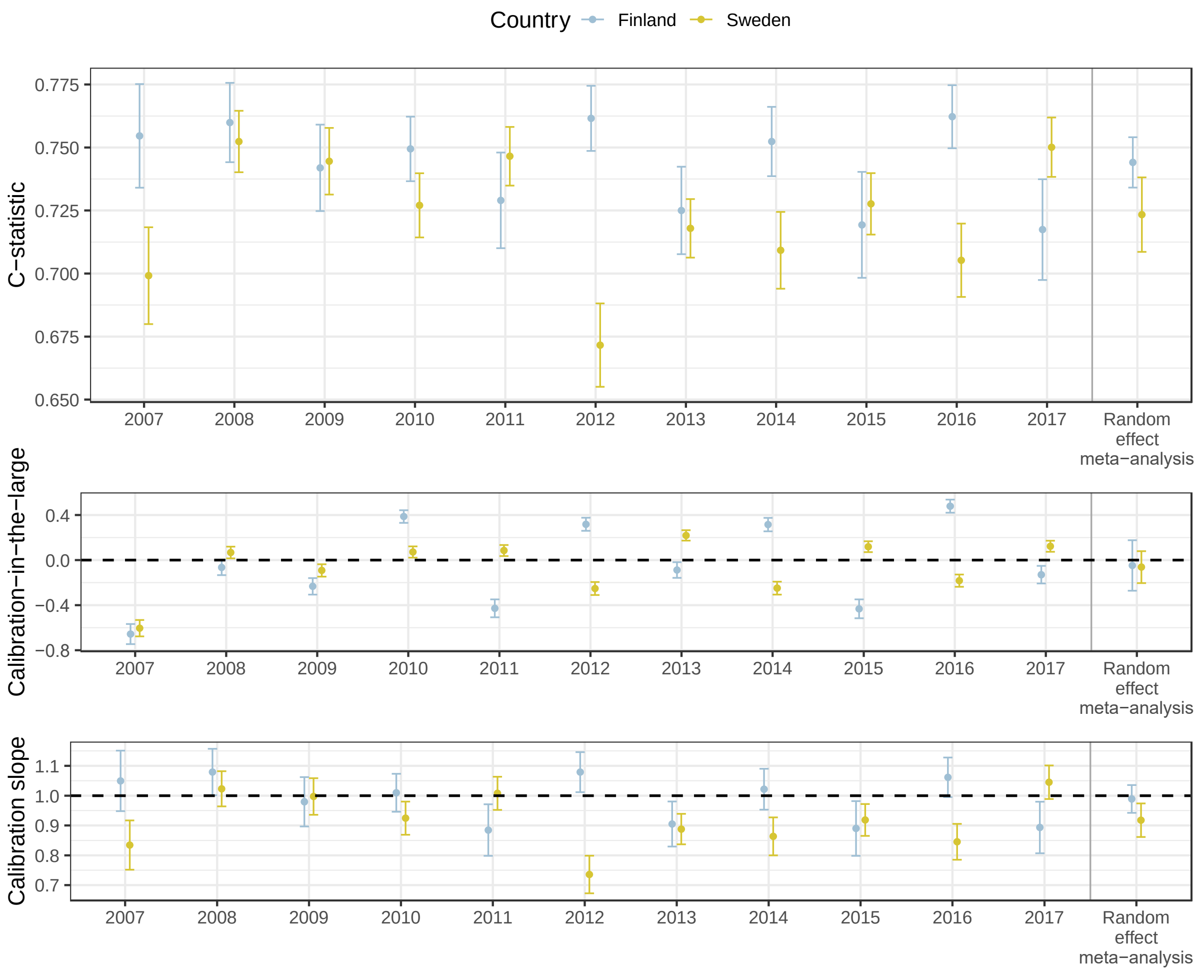

**Supplementary figure 6** The model performance metrics of the clinical prediction model, shown for the years used in the model training in the Finnish data.

The Finnish metrics are obtained from leave-out cross validation, where each epidemic year is kept as a testing data in turn, the model is trained in all other data and tested in the test fold. The Swedish metrics are from the external test data, and no model training is done in the Swedish data. Note that the metrics, especially the c-statistics are not directly comparable, as in the Swedish data, the timing of the RSV epidemic is estimated from the previous years, and in the Finnish data, the actual epidemic timing is used.

The year in the x axis indicates the RSV epidemics; We used the 1st of June as a start for the RSV epidemic year (for example, all children born between the 1st of June 2007-31st of May 2008 were grouped for RSV epidemic year 2018). In calibration metrics, dashed lines indicate perfect calibration. Random-effect meta-analysis is the meta-analysed metric from all shown epidemic years.

Calibration-in-the-large in the complete pooled external validation data was -0.06 (95% CI -0.05 to -0.08), and the calibration slope was 0.94 (95% CI 0.92 - 0.95), likely explained by the slightly lower outcome rate in Sweden.

### 7 Discrimination and calibration according to the outcome prevalence

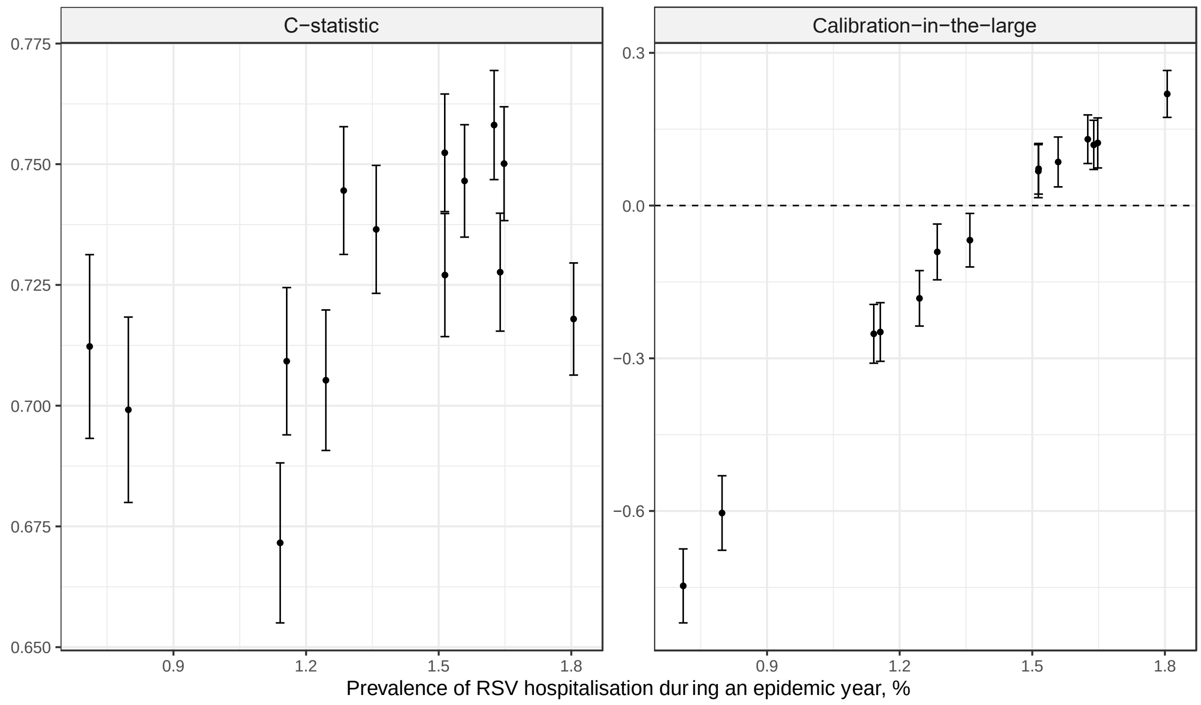

**Supplementary figure 7** The model performance metrics (C-statistic for discrimination and calibration-in-the-large for calibration) shown according to the prevalence of the outcome, i.e. the percentage of infants hospitalised in Sweden during RSV epidemics 2007-2020. Each dot represents an RSV epidemic year, i.e. children born between june-may. X-axis shows the percentage of the children having the outcome, i.e. severe RSV-LRTI requiring hospitalisation. Y-axis shows the respective performance metric. Dashed line in right panel is the reference for perfect calibration-in-the-large.

### 8 The calibration plots

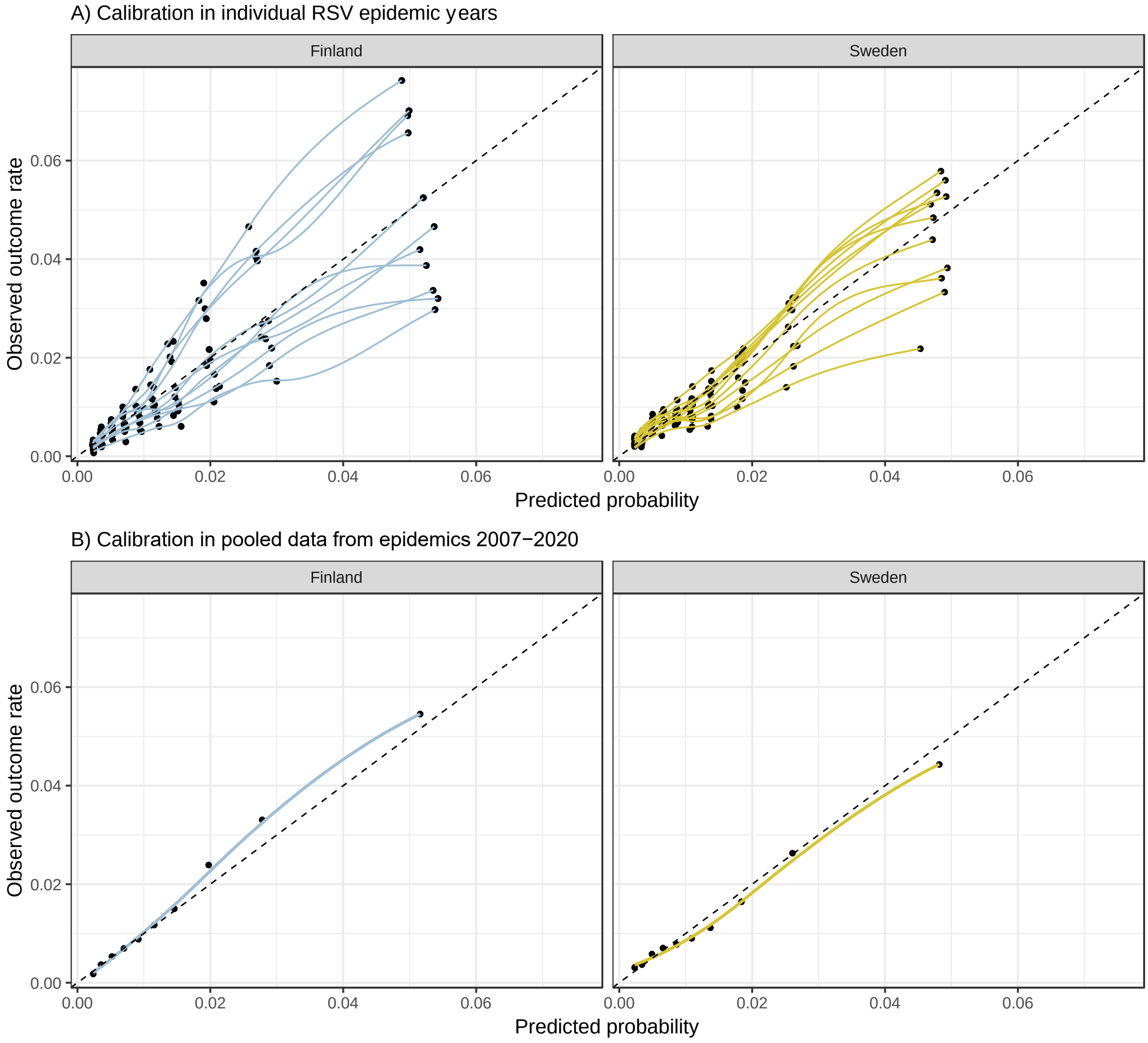

**Supplementary figure 8** The calibration plots, i.e. the predicted vs the observed probabilities in equally sized deciles divided according to the predicted probability, separately for the three epidemic years of this comparison. Each dot represents a decile, and axes show respectively their mean predicted probability and the mean observed outcome rate

In Panel A, The data for RSV epidemics 2007-2017, corresponding to the Finnish training data, are shown individually for each year. We used the 1st of June as a start for the RSV epidemic year (for example, all children born between the 1st of June 2007-31st of May 2008 were grouped for RSV epidemic year 2018). The dashed lines indicate perfect calibration.The Finnish results are obtained from leave-out cross validation, where each epidemic year is kept as a testing data in turn, the model is trained in all other data and tested in the test fold. The Swedish metrics are from the external validation data, and no model training is done in the Swedish data.

In Panel B, calibration data is shown for pooled data covering epidemic years 2007-2020 in both countries. To obtain these pooled results in Finland, we combined the development and validation data and used the final prediction model to assign predicted probabilities to each individual. Similarly to panel A, the Swedish results are from external validation data.

### 9 Comparison of XGboost and clinical prediction model

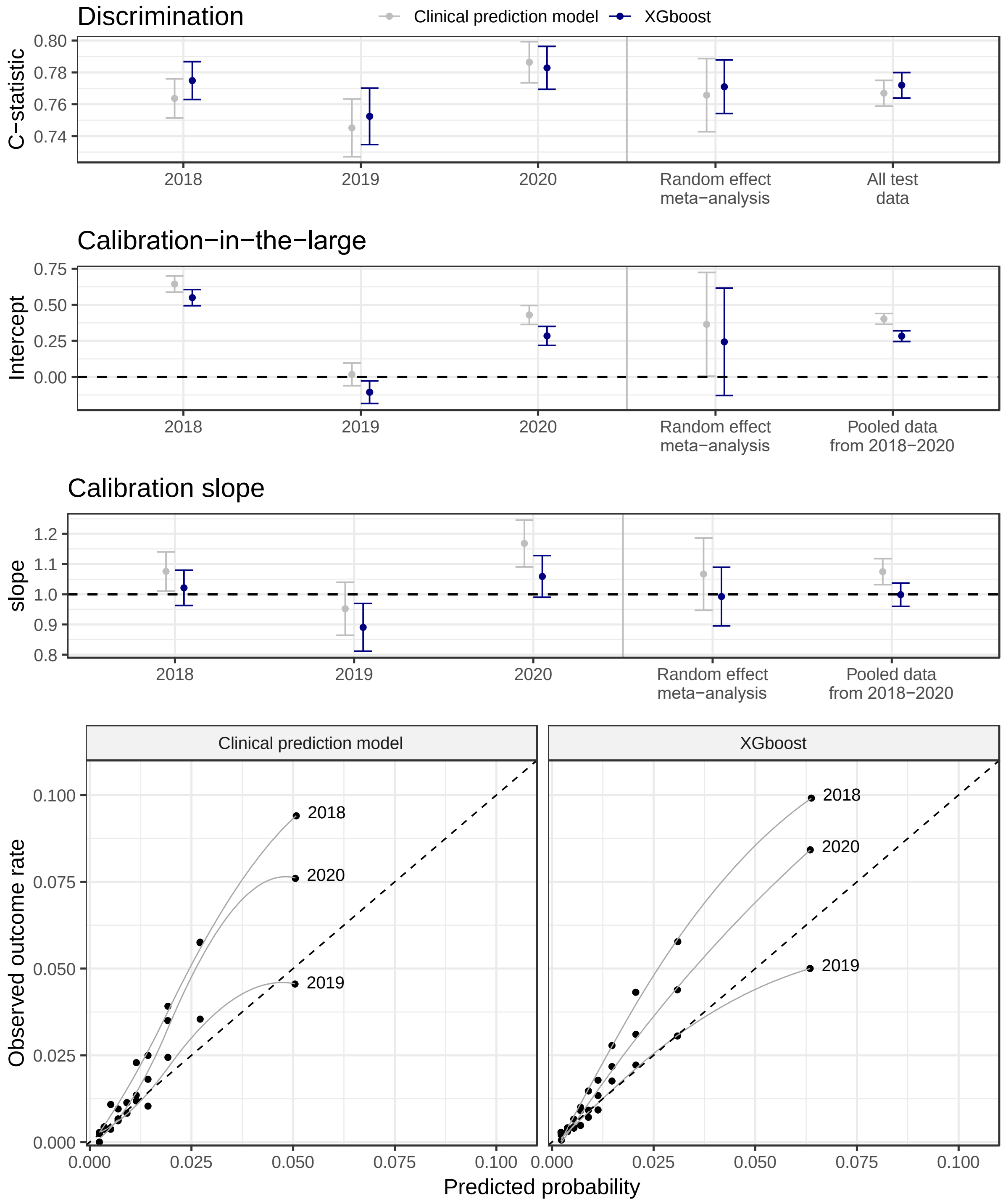

**Supplementary image 9** The discrimination and calibration of the XGboost model shown separately for children predisposed to individual RSV epidemic. The year numbers in the image indicate the RSV epidemics. We used the 1st of June as a start for the RSV epidemic year (for example, all children born between the 1st of June 2018-31st of May 2019 were categorised for RSV epidemic year 2019). To summarise the results of the uppermost 3 panels, the respective metrics from each RSV epidemic are combined with random effect meta-analysis. Also the results obtained from testing the model in the complete held-out test set (all 3 epidemics pooled) are shown for comparison. The 2 lowest panels show the calibration curve, with the test data divided into 10 bins and plotting those bins' mean predicted probability and mean hospitalisation rate against each other. Reference lines show perfect calibration. Note the biennial pattern in the measures, too low predicted probabilities occurring during strong epidemics and lower predicted probabilities in milder epidemics.

### 10 SHAP values of XGboost model

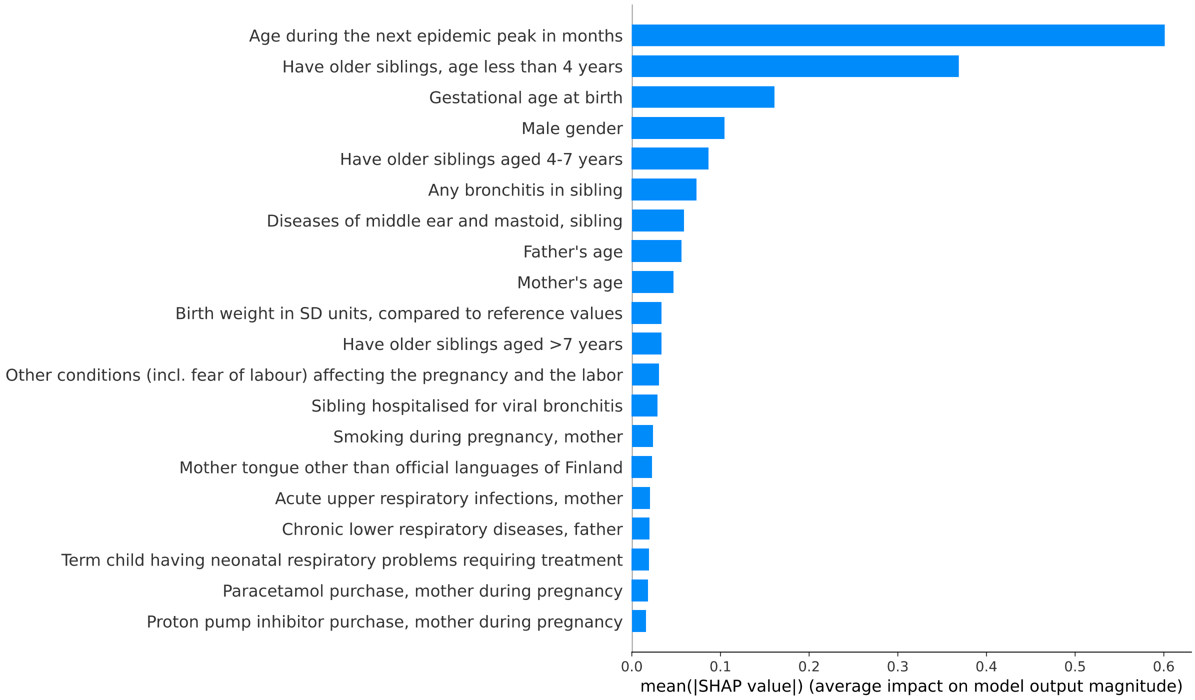

**Supplementary image 10** Variable importance in the XGBoost model for the 20 most important variables, based on the SHapley Additive exPlanation (SHAP) values.[17] Higher SHAP value indicates higher predictor importance for the XGboost model.

### 11 Fairness analysis

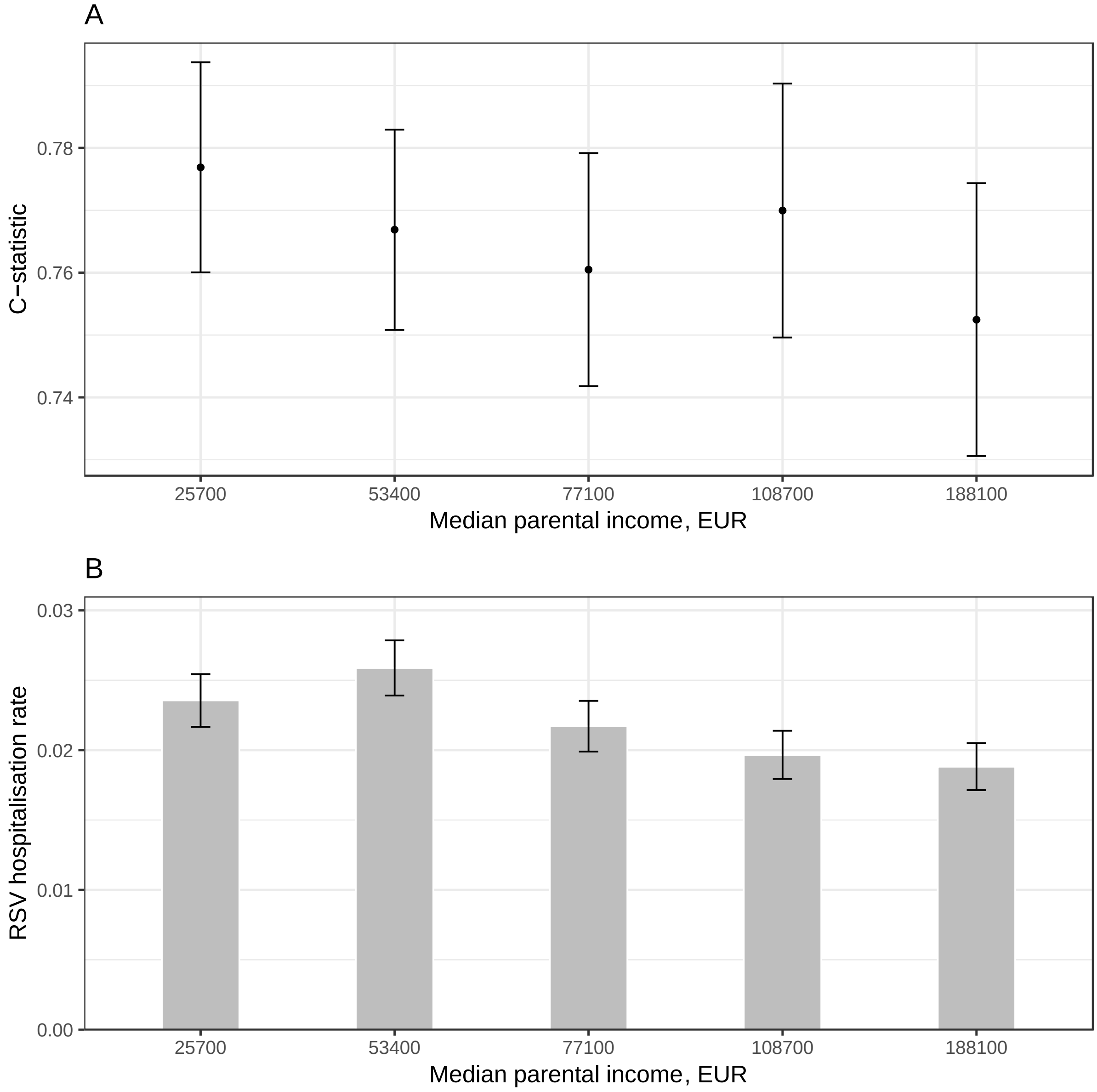

**Supplementary figure 11** The fairness analysis. Panel a) shows the comparison of the C-statistic between parental income quintiles. For descriptive purposes, also the RSV hospitalisation rates for each quintile are shown in panel b). X axis labels show the median income in each quintile and the error bars show 95% confidence intervals of the c-statistic and RSV hospitalisation rate. The results are from Finnish held-out validation data, covering children born between June 2018 and 5/2020. 5962 children (4.8%) In the Finnish internal validation set had missing parental income. In this population, the C-statistic was 0.746 (95% CI 0.710 - 0.781), slightly worse than in all of the quintiles. The C-statistic in the lowest quintile was 0.777 (95% CI 0.760-0.794), compared to 0.753 (0.731-0.775) in the highest quintile (supplementary figure 11) suggesting that the model performed slightly better in, and thus does not harmfully discriminate against children from lower income families.

### REFERENCES

1 Rietveld E, Vergouwe Y, Steyerberg EW, Huysman MWA, de Groot R, Moll HA. Hospitalization for Respiratory Syncytial Virus Infection in Young Children: Development of a Clinical Prediction Rule. *Pediatr Infect Dis J* 2006; **25**: 201–7.

2 Houben ML, Bont L, Wilbrink B, *et al.* Clinical Prediction Rule for RSV Bronchiolitis in Healthy Newborns: Prognostic Birth Cohort Study. *PEDIATRICS* 2011; **127**: 35–41.

3 Blanken MO, Paes B, Anderson EJ, *et al.* Risk scoring tool to predict respiratory syncytial virus hospitalisation in premature infants. *Pediatr Pulmonol* 2018; **53**: 605–12.

4 Blanken MO, Koffijberg H, Nibbelke EE, Rovers MM, Bont L, on behalf of the Dutch RSV Neonatal Network. Prospective Validation of a Prognostic Model for Respiratory Syncytial Virus Bronchiolitis in Late Preterm Infants: A Multicenter Birth Cohort Study. *PLoS ONE* 2013; **8**: e59161.

5 Sampalis JS, Langley J, Carbonell-Estrany X, *et al.* Development and Validation of a Risk Scoring Tool to Predict Respiratory Syncytial Virus Hospitalization in Premature Infants Born at 33 through 35 Completed Weeks of Gestation. *Med Decis Making* 2008; **28**: 471–80.

6 European RSV Risk Factor Study Group, Simões EA, Carbonell-Estrany X, *et al.* A predictive model for respiratory syncytial virus (RSV) hospitalisation of premature infants born at 33–35 weeks of gestational age, based on data from the Spanish FLIP study. *Respir Res* 2008; **9**. DOI:10.1186/1465-9921-9-78.

7 Kurki MI, Karjalainen J, Palta P, *et al.* FinnGen provides genetic insights from a well-phenotyped isolated population. *Nature* 2023; **613**: 508–18.

8 Riley RD, Ensor J, Snell KIE, *et al.* Calculating the sample size required for developing a clinical prediction model. *BMJ* 2020; : m441.

9 Örtqvist AK, Lundholm C, Wettermark B, Ludvigsson JF, Ye W, Almqvist C. Validation of asthma and eczema in population-based Swedish drug and patient registers: VALIDATION OF ASTHMA AND ECZEMA IN SWEDISH REGISTERS. *Pharmacoepidemiol Drug Saf* 2013; **22**: 850–60.

10 Renko M, Tapiainen T. Change in respiratory syncytial virus seasonality in Finland. *Acta Paediatr* 2020; **109**: 202–3.

11 Hamrin J, Bennet R, Berner J, Rotzén-Östlund M, Eriksson M. Rates and risk factors of severe respiratory syncytial virus infection in 2008-2016 compared with 1986-1998. *Acta Paediatr* 2021; **110**: 963–9.

12 Sankilampi U, Hannila M-L, Saari A, Gissler M, Dunkel L. New population-based references for birth weight, length, and head circumference in singletons and twins from 23 to 43 gestation weeks. *Ann Med* 2013; **45**: 446–54.

13 Maršál K, Persson P-H, Larsen T, Lilja H, Selbing A, Sultan B. Intrauterine growth curves based on ultrasonically estimated foetal weights. *Acta Paediatr* 1996; **85**: 843–8.

14 Chaw PS, Wong SWL, Cunningham S, *et al.* Acute Lower Respiratory Infections Associated With Respiratory Syncytial Virus in Children With Underlying Congenital Heart Disease: Systematic Review and Meta-analysis. *J Infect Dis* 2020; **222**: S613–9.

15 Harrell FE. Regression Modeling Strategies: With Applications to Linear Models, Logistic Regression, and Survival Analysis. Springer (New York), 2001.

16 Chen T, Guestrin C. XGBoost: A Scalable Tree Boosting System. In: Proceedings of the 22nd ACM SIGKDD International Conference on Knowledge Discovery and Data Mining. San Francisco California USA: ACM, 2016: 785–94.

17 Lundberg SM, Erion G, Chen H, *et al.* From local explanations to global understanding with explainable AI for trees. *Nat Mach Intell* 2020; **2**: 56–67.
18 Oudega R, Hoes AW, Moons KGM. The Wells Rule Does Not Adequately Rule Out Deep Venous Thrombosis in Primary Care Patients. *Ann Intern Med* 2005; **143**: 100.
19 Rothman KJ, Boice HD, Austin H. Epidemiologic analysis with a programmable calculator, New ed. Boston, Mass: Epidemiology Resources, 1982.
20 Ferguson J, Maturo F, Yusuf S, O'Donnell M. Population attributable fractions for continuously distributed exposures. *Epidemiol Methods* 2020; **9**. DOI:10.1515/em-2019-0037.
21 DeLong ER, DeLong DM, Clarke-Pearson DL. Comparing the Areas under Two or More Correlated Receiver Operating Characteristic Curves: A Nonparametric Approach. *Biometrics* 1988; **44**: 837.
22 American Academy of Pediatrics Committee on Infectious Diseases. Updated Guidance for Palivizumab Prophylaxis Among Infants and Young Children at Increased Risk of Hospitalization for Respiratory Syncytial Virus Infection. *Pediatrics* 2014; **134**: e620–38.
23 Hammitt LL, Dagan R, Yuan Y, *et al.* Nirsevimab for Prevention of RSV in Healthy Late-Preterm and Term Infants. *N Engl J Med* 2022; **386**: 837–46.
24 Griffin MP, Yuan Y, Takas T, *et al.* Single-Dose Nirsevimab for Prevention of RSV in Preterm Infants. *N Engl J Med* 2020; **383**: 415–25.

25 Simões EAF, Madhi SA, Muller WJ, *et al.* Efficacy of nirsevimab against respiratory syncytial virus lower respiratory tract infections in preterm and term infants, and pharmacokinetic extrapolation to infants with congenital heart disease and chronic lung disease: a pooled analysis of randomised controlled trials. *Lancet Child Adolesc Health* 2023; : S2352464222003212.

26 Salisbury D, Ramsay M, Noakes K. Immunisation against infectious disease: the Green book, Updated ed. London: The Stationary Office, 2013.
